# The effect of calcium supplementation in people under 35 years old: A systematic review and meta-analysis of randomized controlled trials

**DOI:** 10.1101/2022.04.14.22273724

**Authors:** Yupeng Liu, Siyu Le, Yi Liu, Huinan Jiang, Binye Ruan, Yufeng Huang, Xuemei Ao, Xudong Shi, Xiaoyi Fu, Shuran Wang

**Affiliations:** Department of Epidemiology and Biostatistics, School of Public Health and Management, Wenzhou Medical University, Wenzhou 325000, China; Department of Nutrition and Food Hygiene, School of Public Health and Management, Wenzhou Medical University, Wenzhou 325000, China

**Keywords:** calcium, peak bone mass, calcium supplementation, osteoporosis

## Abstract

**Background and objective:** The effect of calcium supplementation on bone mineral accretion in people under 35 years old is inconclusive. To comprehensively summarize the evidence for the effect of calcium supplementation on bone mineral accretion in young populations (≤35 years).

**Design:** This is a systematic review and meta-analysis.

**Data sources:** The Pubmed, Embase, ProQuest and CENTRAL databases were systematically searched from database inception to April 25, 2021.

**Eligibility:** Randomized clinical trials assessing the effects of calcium supplementation on bone mineral density (BMD) or bone mineral content (BMC) in people under 35 years old.

**Results:** This systematic review and meta-analysis identified 43 studies involving 7382 subjects. Moderate certainty of evidence showed that calcium supplementation was associated with the accretion of BMD and BMC, especially on femoral neck (SMD 0.627, 95% CI 0.338 to 0.915; SMD 0.364, 95% CI 0.134 to 0.595; respectively) and total body (SMD 0.330, 95% CI 0.163 to 0.496; SMD 0.149, 95% CI 0.006 to 0.291), also with a slight improvement effect on lumbar spine BMC (SMD 0.163, 95% CI 0.008 to 0.317), no effects on total hip BMD and BMC and lumbar spine BMD were observed. Very interestingly, subgroup analyses suggested that the improvement of bone at femoral neck was more pronounced in the peri-PBM population (20-35 years) than the pre-PBM population (<20 years).

**Conclusion:** Our findings provided novel insights and evidence in calcium supplementation, which showed that calcium supplementation significantly improves bone mass, implying that preventive calcium supplementation before or around achieving PBM may be a shift in the window of intervention for osteoporosis.

**Funding:** This work was supported by Wenzhou Medical University grant [89219029].

## Introduction

Osteoporosis is an imperative public health problem, particularly in elderly women.^1-3^ Low bone mass and a fast rate of bone loss at menopause are equal risk factors for future fracture.^4^ A low bone mineral content (BMC) or bone mineral density (BMD) in an elderly person implies a suboptimal bone mass in young adulthood -- related to peak bone mass (PBM), greater bone loss in later life, or both. A number of studies have concluded that increasing calcium intake in older people is unlikely to translate into clinically meaningful reductions in fractures or produce progressive increases in bone mass.^5-8^ It seems that calcium supplementation is meaningless in the elderly. On the other hand, intervention before the achievement of PBM to maximize PBM might have a significant influence on bone health and further prevent osteoporosis later in life. Several clinical trials have shown positive effects of calcium supplementation on BMD or BMC in children.^9 10^ However, several clinical trials have concluded that calcium supplementation may not be associated with calculated bone mass or strength.^11 12^ Narrative reviews have also concluded that calcium supplementation may have small nonprogressive effects on BMD or BMC.^13 14^ To summarize the studies above, there have been considerable debates about whether calcium supplementation has effects on bone health among young people.

Very recently, a study using cross-sectional data from NHANES 2005-2014 concluded that the age at attainment of peak femoral neck BMD, total hip BMD and lumbar spine BMD was 20-24 years old in males and 19-20 years old in females.^15^ Additionally, a plateau is achieved in PBM at approximately 30 years old.^16^ Based on the literature above, we decided to limit the threshold to 35 years old in a conservative manner. Since the results of studies in young people are controversial, we carried out a comprehensive meta-analysis to determine the effectiveness of calcium supplementation for improving BMD or BMC in young people before the age of 35. We also aimed to determine whether any effect would vary by sex, baseline calcium intake, ethnicity, age or sources, duration, and doses of calcium supplementation.

## Methods

This meta-analysis was reported according to Preferred Reporting Items for Systematic Reviews and Meta-analyses guidelines.^17^ The protocol for this meta-analysis is available in PROSPERO (CRD42021251275).

### Literature search

We applied search strategies to the following electronic bibliographic databases: PubMed, EMBASE, ProQuest, and CENTRAL (Cochrane Central Register of Controlled Trials) in April 2021 and updated the search using PubMed in October 2021 for eligible studies addressing the effect of calcium or calcium supplementation, milk or dairy products with BMD or BMC as endpoints. Detailed search strategies are provided in the **Supplementary file 1**. Only randomized controlled trials (RCTs) were included in this study. We also hand-searched conference abstract books. The reference sections and citation lists of the retrieved literature, including original research articles, reviews, editorials, and letters, were reviewed for potentially relevant articles.

### Inclusion criteria

We selected trials based on the following criteria: (1) RCTs comparing calcium or calcium plus vitamin D supplements with a placebo or no treatment; (2) trials involving participants aged under 35 years at baseline; (3) trials providing BMD (g/cm^2^) or BMC (g) data measured by dual energy X ray absorptiometry (DXA) as estimates of bone mass. Exclusion criteria: (1) observational studies, such as cohorts, case–control studies, or cross-sectional studies; (2) participants aged over 35 years; (3) trials of participants who were pregnant or in the lactation period; (4) trials without a placebo or control group; (5) trials supplied with only vitamin D; (6) trials that had essential data missing. Two authors (YPL and SYL) independently screened titles and abstracts, and then full-texts of relevant articles according to the inclusion and exclusion criteria. By thoroughly reading full-texts, the reasons for excluded trials are provided in **Supplementary file 2**.

### Risk-of-bias assessments

The quality of the included RCTs was assessed independently by 2 reviewers (SYL, HNJ) based on the Revised Cochrane risk-of-bias tool for randomized trials (RoB 2),^18^ and each item was graded as low risk, high risk and some concerns. The five domains included the randomization process, deviations from intended interventions, missing outcome data, measurement of the outcome, and selection of the reported result. A general risk conclusion can be drawn from the risk assessment of the above five aspects. We defined the included trials as low quality, high quality and moderate quality based on the overall bias, which is consistent with the RoB 2 tool algorithm. Disagreements were resolved by consensus.

### Data extraction and synthesis

Two researchers (YPL, SYL) independently used a structured data sheet to extract the following information from each study: authors, publication year, participant characteristics, doses of the supplements, baseline dietary calcium intake, duration of trials and follow-up. The absolute changes in BMD or BMC at the lumbar spine, femoral neck, total hip and total body were the primary outcomes we extracted. We categorized the studies into two groups by duration: <18 months and ≥18 months. For studies that presented the percentage change rather than absolute data, we calculated the absolute change value using baseline data, and the standard deviation and percentage change from baseline were consistent with the approach described in the Cochrane Handbook.^18^ If there was missing information, we contacted the corresponding author and obtained the data. (If no reply was received for over three months, we would exclude the article.)

### Statistical analysis

The association of calcium with or without vitamin D supplements with BMD and BMC was assessed. We pooled the data (study-level) from each study using random-effects models in a conservative manner. The standardized mean difference (SMD) and corresponding 95% confidence intervals (CIs) were reported. We performed predesigned subgroup analyses based on the following aspects: sex (female vs. male) and age at baseline (<20 vs. ≥20 years, representing the prepeak and peripeak subgroups, respectively; all analysed trials were divided into two groups by the age of achieving PBM (determined as 20 years old), regions (Asian and Western), sources of calcium supplementation (dietary vs. calcium supplements), and bias risk of each individual trial. We further conducted some post hoc subgroup analyses according to the level of calcium intake at baseline (< 714 vs. ≥ 714 mg/day, based on the median value) and the calcium supplementation dose (<1000 versus ≥1000 mg/day, based on the median value). To assess how long the beneficial effect would be maintained, we performed post hoc subgroup analyses according to the duration, taking into account different calcium supplementation periods and different follow-up periods across the trials. Sensitivity analyses included evaluations using fixed-effect models and excluding low-quality trials. In these aforementioned subgroup analyses, if the number of eligible studies in subgroups was less than three, we conducted a sensitivity analysis by excluding the subgroup with fewer than three studies. An effect size of ≥ 0.20 and < 0.50 was considered small, ≥ 0.50 and < 0.80 was considered medium, and ≥ 0.80 was considered large using Cohen’s criteria.^19^

We assessed heterogeneity between studies using the I^2^ statistic. We performed meta-regression for sample size, age, sex, and supplementation differences to explain the heterogeneity between studies. We performed cumulative meta-analyses based on the sample size to compare with the primary outcomes. We assessed publication bias by examining funnel plots when the number of trials was 10 or more and used Begg’s rank correlation and Egger’s linear regression tests.^20^ Furthermore, we robustly adjusted for the summarized results by applying Duval and Tweedie’s trim and fill method^21^ All analyses were performed by Comprehensive Meta Analysis (version 2.0, Biostat, Englewood, NJ). All tests were 2-tailed, and P< .05 was considered statistically significant. Two reviewers (SYL, YL) independently applied the Grading of Recommendations Assessment, Development and Evaluation (GRADE) system to assess the overall quality of evidence. The quality of evidence for each outcome was classified as either high, moderate, low, or very low based on the evaluation for study design, bias risk, inconsistency, indirectness, imprecision, publication bias, and confounding bias. GRADE pro version 3.6 was used to grade the overall quality of evidence and prepare the summary-of-findings table. Every decision to downgrade or upgrade the studies was labeled using footnotes. Any disagreements were resolved by consensus.

## Results

### Study characteristics

Of the 5518 references screened, we identified 43 eligible RCTs (**Figure 1**) involving 7382 subjects.^9-12 22-60^ **Table 1** shows the baseline characteristics of the included studies. Of the 43 RCTs, 20 used dietary sources of calcium,^11 12 22 23 25 27-29 40 43-46 48 49 53 54 56 60^ and 23 used calcium supplements (including calcium, calcium citrate malate and calcium phosphate).^9 10 24 26 30-39 41 42 47 50-52 55 57 59^ The median baseline dietary calcium intake was 714 mg/day; the duration of calcium supplementation intervention did not exceed 2 years in most trials (38/43); and the dose of calcium intervention did not exceed 1000 mg/day in most trials (38/43). Of all the included trials, 23 trials were categorized as low risk of bias; 16, as moderate risk; and 4, as high risk (**Supplementary file 3**).

**Table 1.**
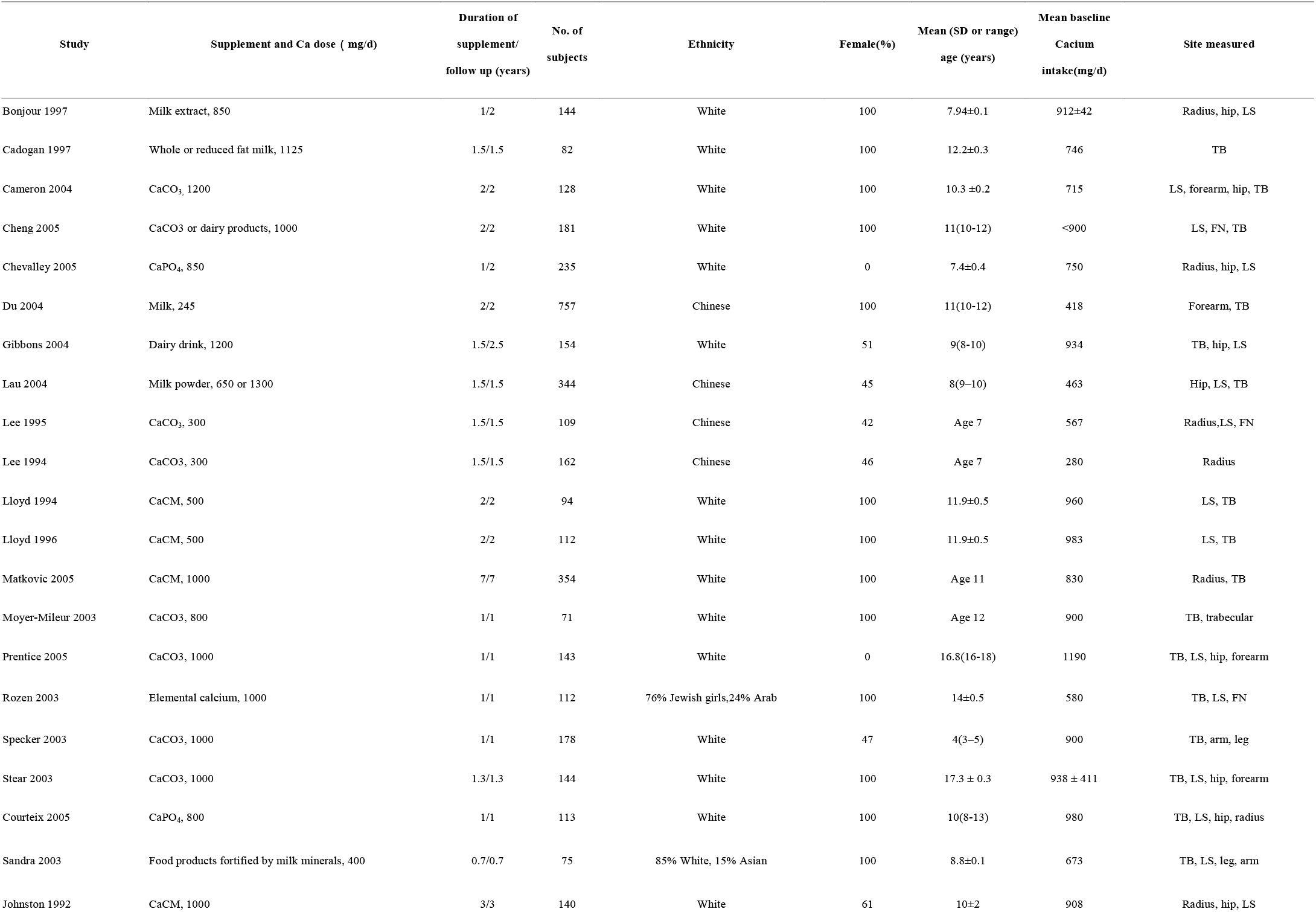
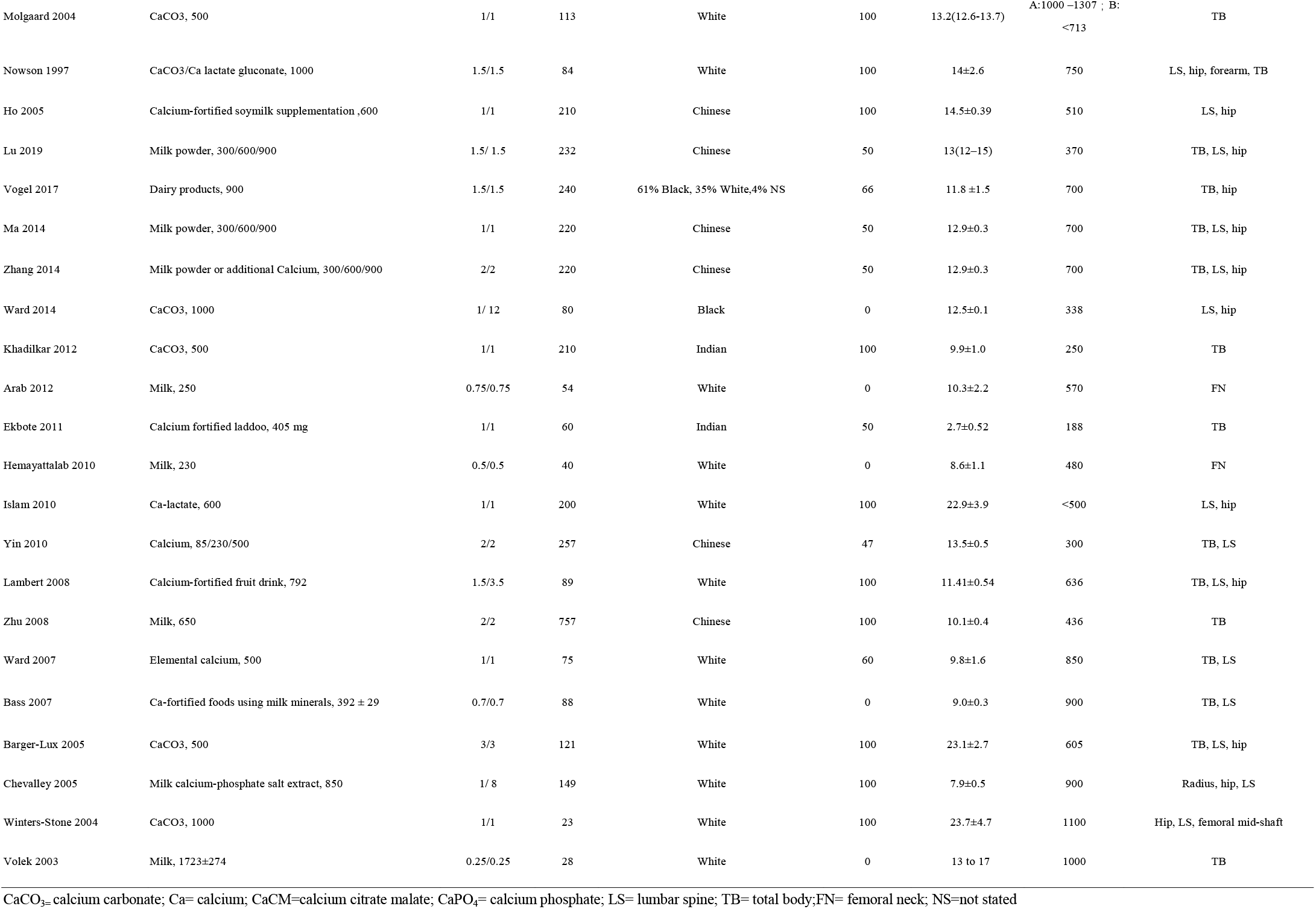
Characteristics of included studies.

**Figure 1:**
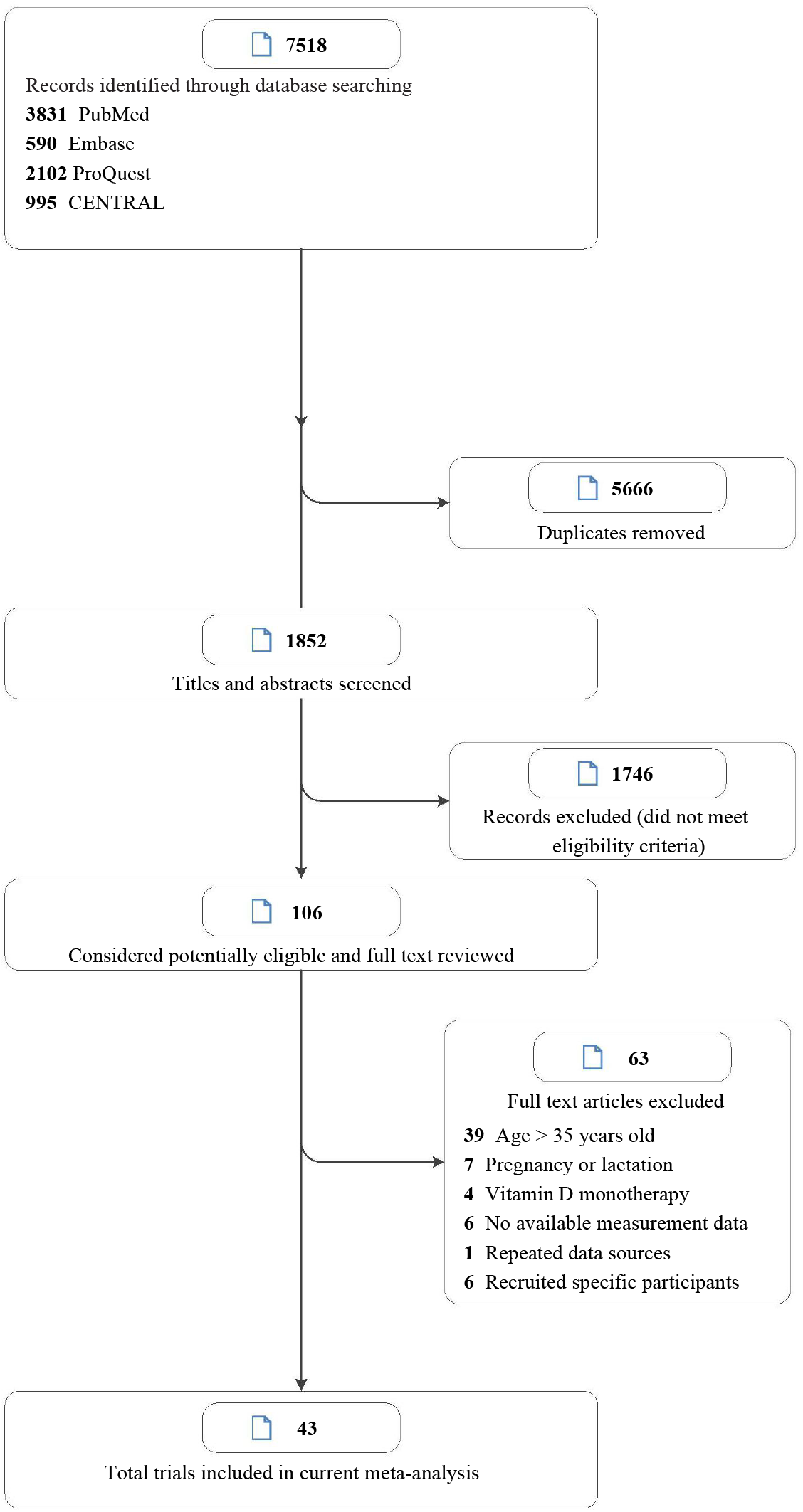
Study selection.

### Primary analyses

**Figure 2, Figure 2-figure supplement 1, Figure 3** and **Figure 3-figure supplement 1** shows the summarized effect estimates. For total body, moderate evidence showed that calcium supplementation significantly improved BMD levels with an SMD of 0.330 (95% CI: 0.163 to 0.496, P<.001) and slightly improved BMC levels with an SMD of 0.149 (95%CI: 0.006 to 0.291, P<.001). At the femoral neck, we found a stronger and moderate protective effect on BMD (0.627, 95% CI: 0.338 to 0.915, P<.001) and a small improvement effect on BMC (0.364, 95% CI: 0.134 to 0.595, P=0.002). Meanwhile, a slight but significant improvement in BMC was observed for the lumbar spine (0.163, 95% CI: 0.008 to 0.317, P=0.039). However, calcium supplementation did not improve the BMD levels at the lumbar spine (0.090, 95% CI: -0.044 to 0.224, P=0.190) or total hip (0.257, 95% CI: -0.053 to 0.566, P=0.104) or the BMC level at the total hip (0.116, 95% CI: -0.382 to 0.614, P=0.648).

**Figure 2.**
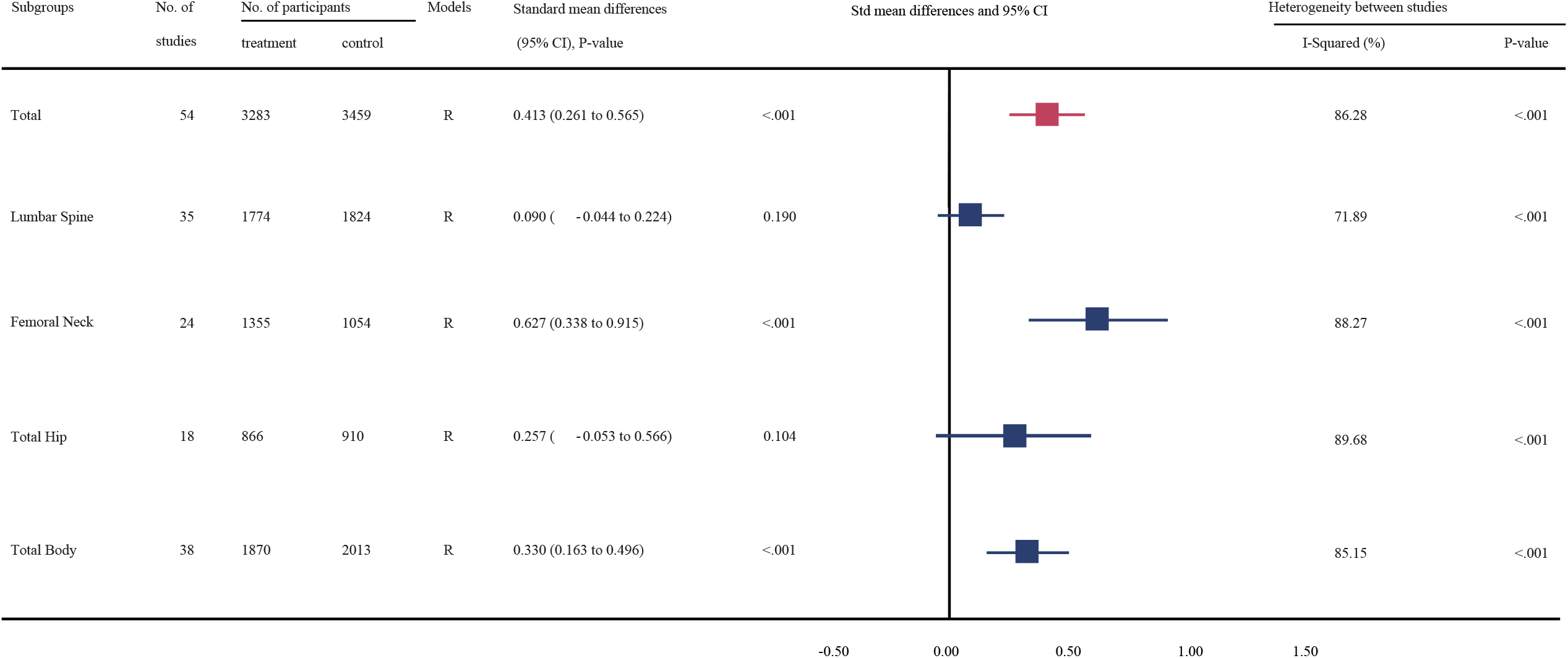
Effect of calcium supplementation on bone mineral density (BMD) in each sites.

**Figure 3.**
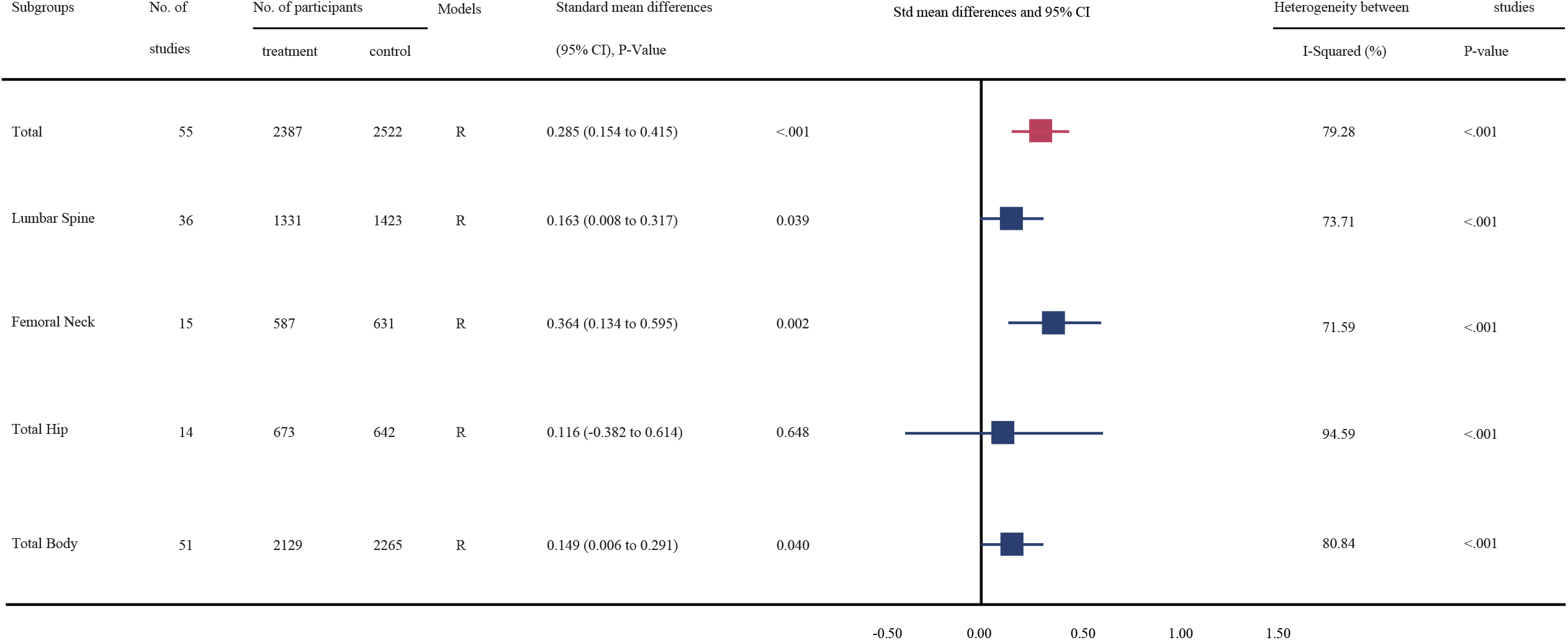
Effect of calcium supplementation on bone mineral content (BMC) in each sites.

### Subgroup analyses

**Table2** and **3** shows the results of subgroup analyses. To explore whether the observed effect differed by the age of participants, we divided these participants into two subgroups: prepeak (< 20 years) and peripeak (≥20-35 years), and the results were generally consistent with the findings from the primary analyses. Notably, the improvement effect on both BMD and BMC at the femoral neck (see **Figure 4**) tended to be stronger in the peripeak subjects than in the prepeak subjects (0.852, 95% CI: 0.257 to 1.446 vs. 0.600, 95% CI: 0.292 to 0.909 [for BMD] and 1.045, 95% CI: 0.701 to 1.39 vs. 0.249, 95% CI: 0.043 to 0.454 [for BMC], respectively).

**Figure 4.**
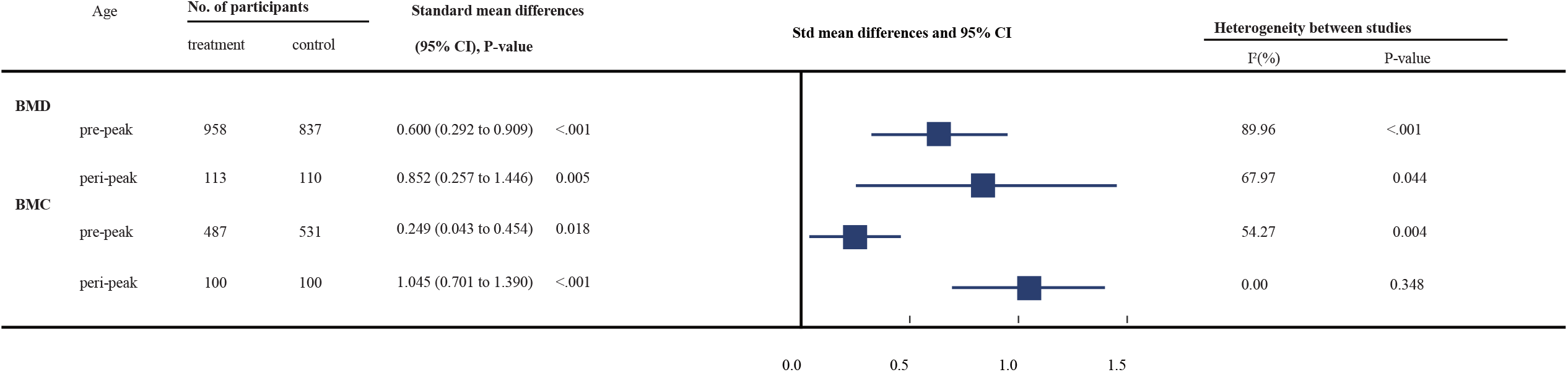
Comparison of the effect of calcium supplementation between pre-peak and peri-peak participants.

Subgroup analyses by the duration of calcium supplementation showed that the improvement effects on both BMD and BMC of the femoral neck were stronger in the subgroup with <18 months than in the subgroup with ≥18 months. However, regarding total body BMD, the effect of calcium supplementation in the subgroup with ≥ 18 months duration was slightly greater than that in the other subgroup.

Regarding the sex of subjects, we found a stronger beneficial effect on femoral neck BMD and BMC in women-only trials (0.712, 95% CI: 0.149 to 1.275, P=0.013; 0.742, 95% CI: 0.267 to 1.217, P=0.002, respectively) than in trials including men and women (0.556, 95% CI: 0.233 to 0.879, P=0.001; 0.195, 95% CI: -0.027 to 0.418, P=0.086).

When considering the sources of participants, the improvement effects on femoral neck and total body BMD or on femoral neck and lumbar spine BMC were obviously stronger in Western countries than in Asian countries.

Subgroup analyses by the level of dietary calcium intake at baseline showed that, for femoral neck BMD, the beneficial effect was significant only in the lower subgroup receiving <714 mg/day (0.581, 95% CI: 0.266 to 0.896; P<.001); for total body BMD, the beneficial effect was slightly greater in the lower subgroup receiving <714 mg/day (0.363, 95% CI: 0.127 to 0.599; P=0.003); for total hip BMD and lumbar spine BMC, however, the beneficial effects were statistically significant in the higher subgroup receiving ≥714 mg/day (0.723, 95% CI: 0.245 to 1.201; P=0.003 and 0.2, 95% CI: 0.052 to 0.348; P=0.008, respectively).

Subgroup analyses based on calcium supplement dosages demonstrated a statistically significant effect on femoral neck and total body BMD in the lower dose subgroup receiving <1000 mg/day (0.717, 95% CI: 0.349 to 1.085; P<.001 and 0.392, 95% CI: 0.161 to 0.624; P=0.001, respectively) but not in the higher dose subgroup receiving ≥1000 mg/day.

When considering the different sources of calcium, both calcium sources from dietary intake and additional calcium supplements exerted significantly positive effects on femoral neck BMD (0.728, 95% CI: 0.311 to 1.144, P<.001; 0.510, 95% CI: 0.101 to 0.919, P=0.014) and total body BMD (0.290, 95% CI: 0.054 to 0.526, P=0.016; 0.405, 95% CI: 0.195 to 0.615, P<.001). For BMCs of the lumbar spine and femoral neck, only calcium supplements other than dietary intake had a significant improvement effect.

To explore the longevity of the beneficial effect, we performed subgroup analyses and found that calcium supplementation improved the BMD levels during the follow-up periods after the end of intervention, and the beneficial effect was maintained for at least 1 year after the intervention (0.933, 95% CI: 0.323 to 1.664, P=0.004). However, this beneficial effect seemed to disappear when the follow-up period exceeded 2 years.

### Sensitivity analysis

Sensitivity analyses including only trials with a low risk of bias (high quality, see **Supplementary file 4**) showed that the improvement effects on femoral neck BMD and BMC remained statistically significant and stable (0.356, 95% CI: 0.064 to 0.648, P=0.017; 0.249, 95% CI: 0.043 to 0.454, P=0.018). The result for total body BMD was also stable (0.343, 95% CI: 0.098 to 0.588, P=0.006). However, for lumbar spine and total body BMCs, the positive effect was not statistically significant. For other sites, the results were generally consistent with those of the primary analyses. Additional sensitivity analyses using fixed-effect models (see **Supplementary file 5**), performing cumulative meta-analysis (see **Supplementary file 6**) and excluding studies had been included in previous meta-analysis (see **Supplementary file 7**) showed generally consistent results with the primary analyses.

### GRADE scoring

**Supplementary file 8** shows a summary of the GRADE assessments of the overall certainty of the evidence for the effect of calcium supplementation on bone measurements. The evidence was graded as moderate for all sites. All of these outcomes were downgraded for inconsistency. For femoral neck BMD, it was downgraded because of strongly suspected publication bias, however, it was upgraded due to the effect size was over 0.5. In summary, the outcome of femoral neck BMD was graded as moderate.

### Heterogeneity analysis

In general, the heterogeneity between trials was obvious in the analysis for BMD (P<.001, I^2^=86.28%) and slightly smaller for BMC (P<.001, I^2^=79.28%). The intertrial heterogeneity was significantly distinct across the sites measured. Subgroup analyses and meta-regression analyses suggested that this heterogeneity could be explained partially by differences in baseline calcium intake levels, sex and region of participants (**Table 2, 3** and **Supplementary file 9**).

**Table 2.** **Subgroup analysis of bone mineral density (BMD) between calcium supplementation and control for each variable at lumbar spine, femoral neck, total hip and total body**

| Variable | No. of datasets | No. of participants | BMD difference(95% CI), P Value | Heterogeneity between |  | P Value<sup>a</sup> |
| --- | --- | --- | --- | --- | --- | --- |
|  |  |  |  | studies |  |  |
|  |  |  |  | I<sup>2</sup> (%) | P-value |  |
| Lumbar Spine |  |  |  |  |  |  |
| age |  |  |  |  |  |  |
| pre-peak | 31 | 3104 | 0.093 (-0.047 to 0.233), 0.192 | 71.54 | <.001 | 0.866 |
| peri-peak | 4 | 344 | 0.078 (-0.471 to 0.627), 0.780 | 79.82 | 0.002 |  |
| duration |  |  |  |  |  |  |
| <18 months | 14 | 1420 | 0.066 (-0.069 to 0.202), 0.335 | 32.75 | 0.113 | 0.905 |
| ≥18 months | 21 | 2178 | 0.106 (-0.104 to 0.316), 0.322 | 80.31 | <.001 |  |
| sex |  |  |  |  |  |  |
| women-only trials | 13 | 1466 | 0.36 (0.067 to 0.653), 0.016 | 83.71 | <.001 | 0.011 |
| trials with men and women | 22 | 2181 | -0.057 (-0.162 to 0.048), 0.284 | 27.53 | 0.115 |  |
| regions |  |  |  |  |  |  |
| Asian | 18 | 1492 | -0.012 (-0.117 to -0.094), 0.829 | 12.70 | 0.302 | 0.177 |
| Western | 17 | 1956 | 0.222 (-0.03 to 0.473), 0.084 | 83.62 | <.001 |  |
| baseline calcium intake, mg/d |  |  |  |  |  |  |
| <714 | 23 | 2014 | 0.062 (-0.109 to 0.234), 0.477 | 73.19 | <.001 | 0.561 |
| ≥714 | 12 | 1434 | 0.145 (-0.080 to 0.370), 0.207 | 71.17 | <.001 |  |
| calcium dose, mg/d |  |  |  |  |  |  |
| <1000 | 26 | 2172 | 0.103 (-0.062 to 0.269), 0.222 | 75.30 | <.001 | 0.806 |
| ≥1000 | 9 | 1056 | 0.050 (-0.177 to 0.276), 0.667 | 59.22 | 0.012 |  |
| types of calcium supplement |  |  |  |  |  |  |
| dietary calcium | 18 | 1690 | 0.104 (-0.104 to 0.311), 0.328 | 77.83 | <.001 | 0.870 |
| calcium supplementation | 17 | 1758 | 0.075(-0.099-0.249), 0.396 | 63.66 | <.001 |  |
| Femoral Neck |  |  |  |  |  |  |
| age |  |  |  |  |  |  |
| pre-peak | 21 | 1795 | 0.600 (0.292 to 0.909), <.001 | 88.68 | <.001 | 0.138 |
| peri-peak | 3 | 223 | 0.852 (0.257 to 1.446), 0.005 | 67.97 | 0.044 |  |
| duration |  |  |  |  |  |  |
| <18 months | 15 | 1457 | 0.824 (0.383 to 1.266), <.001 | 91.06 | <.001 | 0.578 |
| ≥18 months | 9 | 952 | 0.378 (0.047 to 0.709), 0.025 | 79.12 | <.001 |  |
| sex |  |  |  |  |  |  |
| women-only trials | 8 | 840 | 0.712 (0.149 to 1.275), 0.013 | 90.89 | <.001 | 0.963 |
| trials with men and women | 16 | 1262 | 0.560 (0.233 to 0.879), 0.001 | 85.41 | <.001 |  |
| regions |  |  |  |  |  |  |
| Asian | 10 | 793 | 0.091 (-0.047 to 0.230), 0.197 | 0.00 | 0.441 | 0.115 |
| Western | 14 | 1309 | 1.078 (0.603 to 1.552), <.001 | 91.53 | <.001 |  |
| baseline calcium intake, mg/d |  |  |  |  |  |  |
| <714 | 17 | 1159 | 0.581 (0.266 to 0.896), <.001 | 84.10 | <.001 | 0.57 |

**Table 2. Subgroup analysis of bone mineral density (BMD) between calcium supplementation and control for each variable at lumbar spine, femoral neck, total hip and total body (continued)**
|  |  |  |  |  |  |  |
| --- | --- | --- | --- | --- | --- | --- |
| ≥714 | 7 | 903 | 0.680 (0.036 to 1.323), 0.038 | 93.43 | <.001 |  |
| calcium dose, mg/d |  |  |  |  |  |  |
| <1000 | 18 | 1371 | 0.717 (0.349 to 1.085), <.001 | 89.52 | <.001 | 0.488 |
| ≥1000 | 6 | 731 | 0.421 (-0.055 to 0.897), 0.083 | 85.12 | <.001 |  |
| types of calcium supplement |  |  |  |  |  |  |
| dietary calcium | 15 | 1071 | 0.728 (0.311 to 1.144), 0.001 | 89.73 | <.001 | 0.635 |
| calcium supplementation | 9 | 1031 | 0.510 (0.101 to 0.919), 0.014 | 86.60 | <.001 |  |
| Total Hip |  |  |  |  |  |  |
| age |  |  |  |  |  |  |
| pre-peak | 16 | 1539 | 0.336 (0.031 to 0.642), 0.031 | 88.43 | <.001 | 0.119 |
| peri-peak | 2 | 144 | -0.465 (-1.409 to 0.479), 0.334 | 77.90 | 0.033 |  |
| duration |  |  |  |  |  |  |
| <18 months | 6 | 485 | 0.076 (-0.102 to 0.255), 0.402 | 0.00 | 0.963 | 0.935 |
| ≥18 months | 12 | 1291 | 0.351 (-0.102 to 0.805), 0.129 | 93.24 | <.001 |  |
| sex |  |  |  |  |  |  |
| women-only trials | 5 | 527 | 0.483 (-0.479 to 1.444), 0.325 | 95.75 | 0.000 | 0.932 |
| trials with men and women | 13 | 1070 | 0.181 (-0.103 to 0.465), 0.211 | 83.03 | 0.000 |  |
| regions |  |  |  |  |  |  |
| Asian | 13 | 1126 | 0.096 (-0.127 to 0.319), 0.399 | 73.92 | 0.000 | 0.579 |
| Western | 5 | 471 | 0.690 (-0.429 to 1.81), 0.227 | 96.33 | 0.000 |  |
| baseline calcium intake, mg/d |  |  |  |  |  |  |
| <714 | 15 | 1336 | 0.179 (-0.148 to 0.507), 0.283 | 89.55 | 0.000 | 0.023 |
| ≥714 | 3 | 261 | 0.723 (0.245 to 1.201), 0.003 | 60.02 | 0.082 |  |
| calcium dose, mg/d |  |  |  |  |  |  |
| <1000 | 14 | 1092 | 0.189 (-0.179 to 0.557), 0.314 | 90.28 | 0.000 | 0.329 |
| ≥1000 | 4 | 505 | 0.513 (-0.024 to 1.05), 0.061 | 84.04 | 0.000 |  |
| types of calcium supplement |  |  |  |  |  |  |
| dietary calcium | 15 | 1369 | 0.314 (-0.006 to 0.634), 0.054 | 88.89 | 0.000 | 0.421 |
| calcium supplementation | 3 | 228 | -0.046 (-1.148 to 1.056), 0.935 | 92.84 | 0.000 |  |
| Total Body |  |  |  |  |  |  |
| age |  |  |  |  |  |  |
| pre-peak | 38 | 3883 | 0.330 (0.163 to 0.496), <.001 | 85.15 | <.001 | .. |
| peri-peak | .. | .. | .. | .. | .. |  |
| duration |  |  |  |  |  |  |
| <18 months | 12 | 986 | 0.324 (0.035 to 0.614), 0.028 | 79.55 | <.001 | 0.775 |
| ≥18 months | 26 | 2897 | 0.334 (0.129 to 0.539), 0.001 | 87.15 | <.001 |  |
| sex |  |  |  |  |  |  |
| women-only trials | 18 | 2359 | 0.569 (0.328 to 0.810), <.001 | 87.66 | <.001 | 0.036 |
| trials with men and women | 20 | 1558 | 0.104 (-0.089 to 0.296), 0.292 | 73.86 | <.001 |  |

**Table 2. Subgroup analysis of bone mineral density (BMD) between calcium supplementation and control for each variable at lumbar spine, femoral neck, total hip and total body (continued)**
|  |  |  |  |  |  |  |
| --- | --- | --- | --- | --- | --- | --- |
| ethnicity |  |  |  |  |  |  |
| Asian | 23 | 2008 | 0.274 (0.062 to 0.486), 0.011 | 85.67 | <.001 | 0.544 |
| Western | 15 | 1469 | 0.422 (0.143 to 0.701), 0.003 | 85.28 | <.001 |  |
| baseline calcium intake, mg/d |  |  |  |  |  |  |
| <714 | 26 | 2356 | 0.363 (0.127 to 0.599), 0.003 | 89.23 | <.001 | 0.140 |
| ≥714 | 12 | 1215 | 0.265 (0.136 to 0.394), <.001 | 22.28 | 0.225 |  |
| calcium dose, mg/d |  |  |  |  |  |  |
| <1000 | 27 | 2612 | 0.392 (0.161 to 0.624), 0.001 | 88.51 | <.001 | 0.484 |
| ≥1000 | 11 | 1285 | 0.189 (0.073 to 0.306), 0.001 | 11.81 | 0.332 |  |
| types of calcium supplement |  |  |  |  |  |  |
| dietary calcium | 24 | 2453 | 0.290 (0.054 to 0.526), 0.016 | 88.33 | <.001 | 0.129 |
| calcium supplementation | 14 | 1464 | 0.405 (0.195 to 0.615), <.001 | 74.22 | <.001 |  |

<sup>a</sup> P value for heterogeneity between subgroups.

**Table 3.**
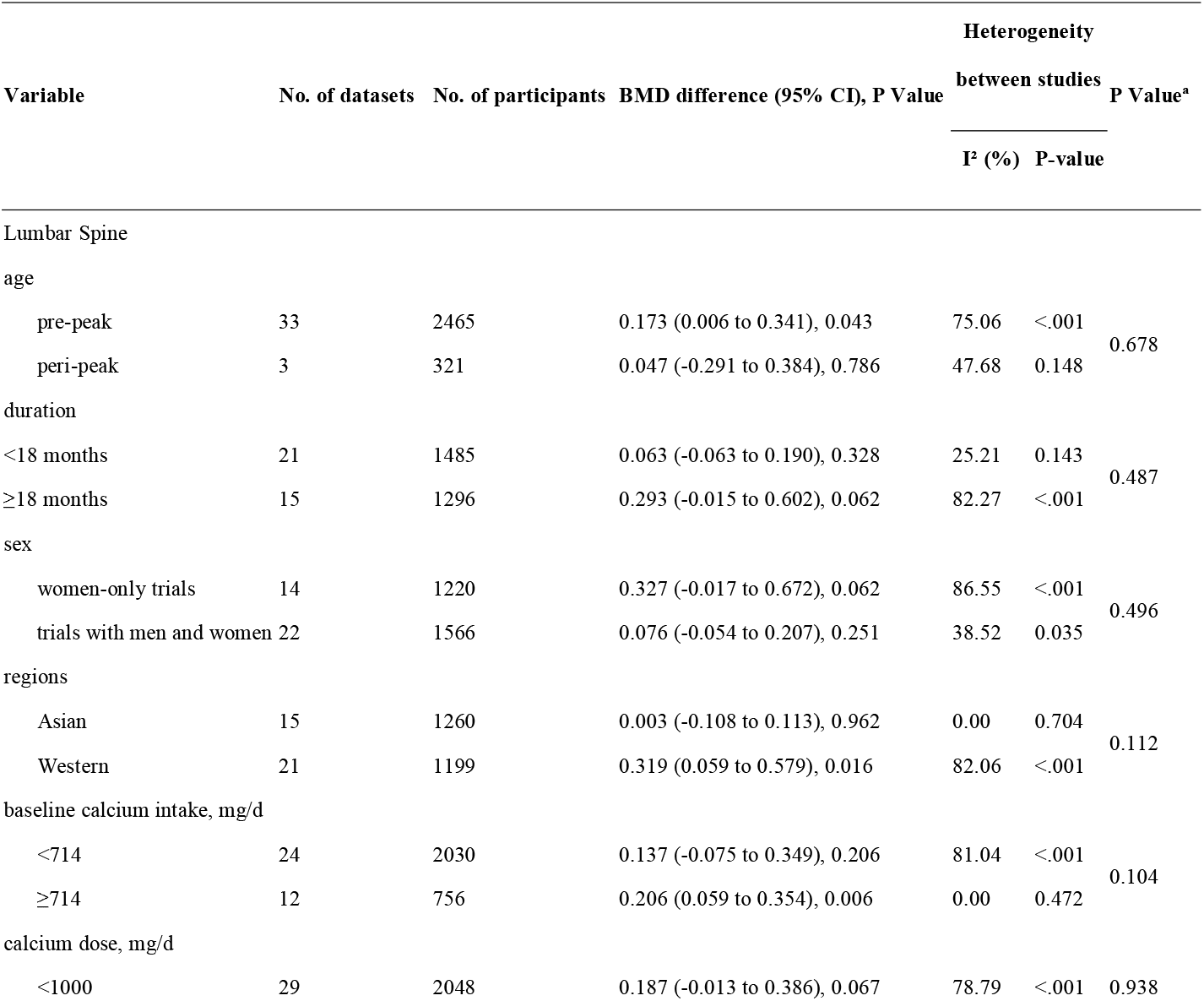
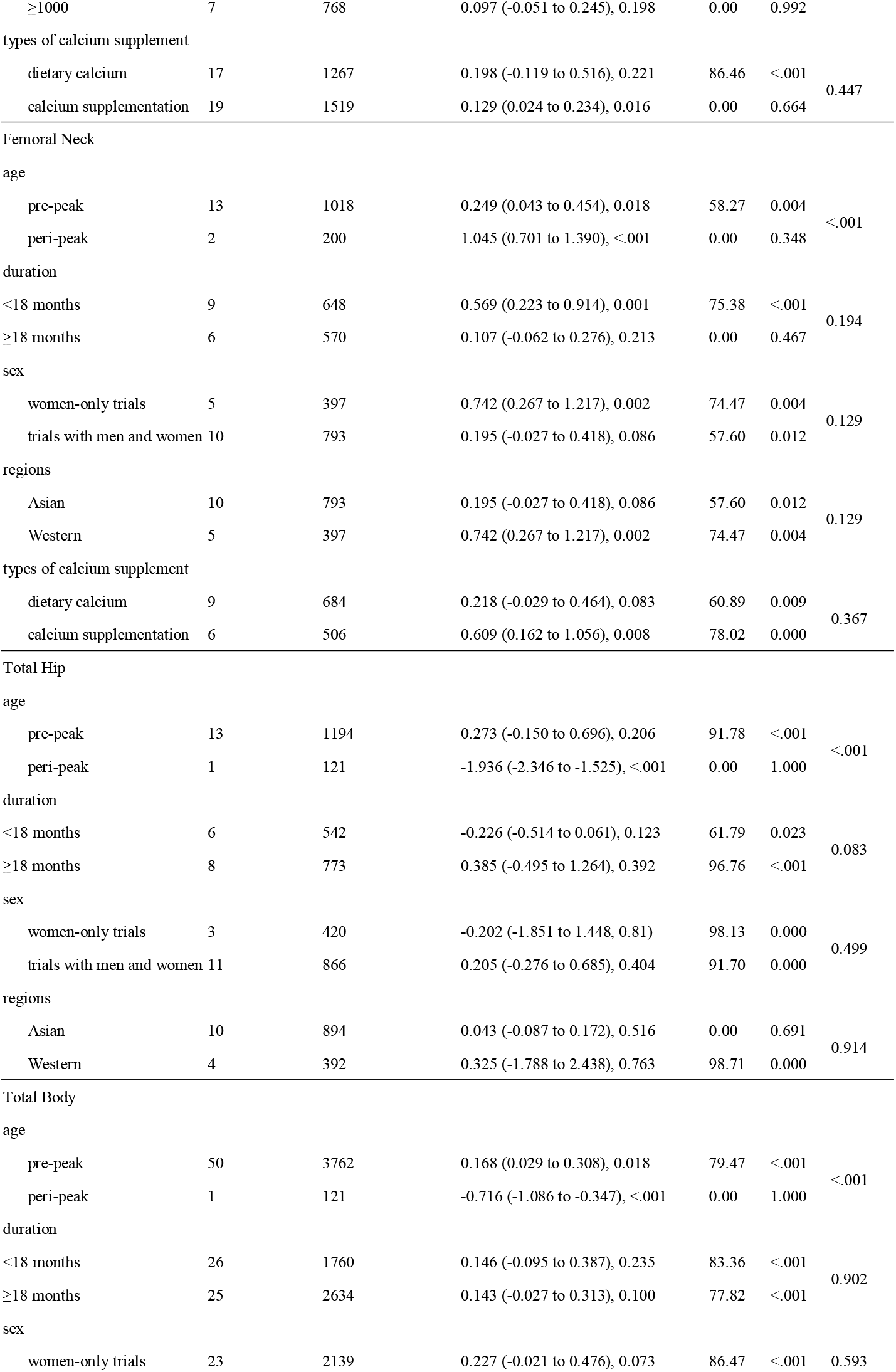
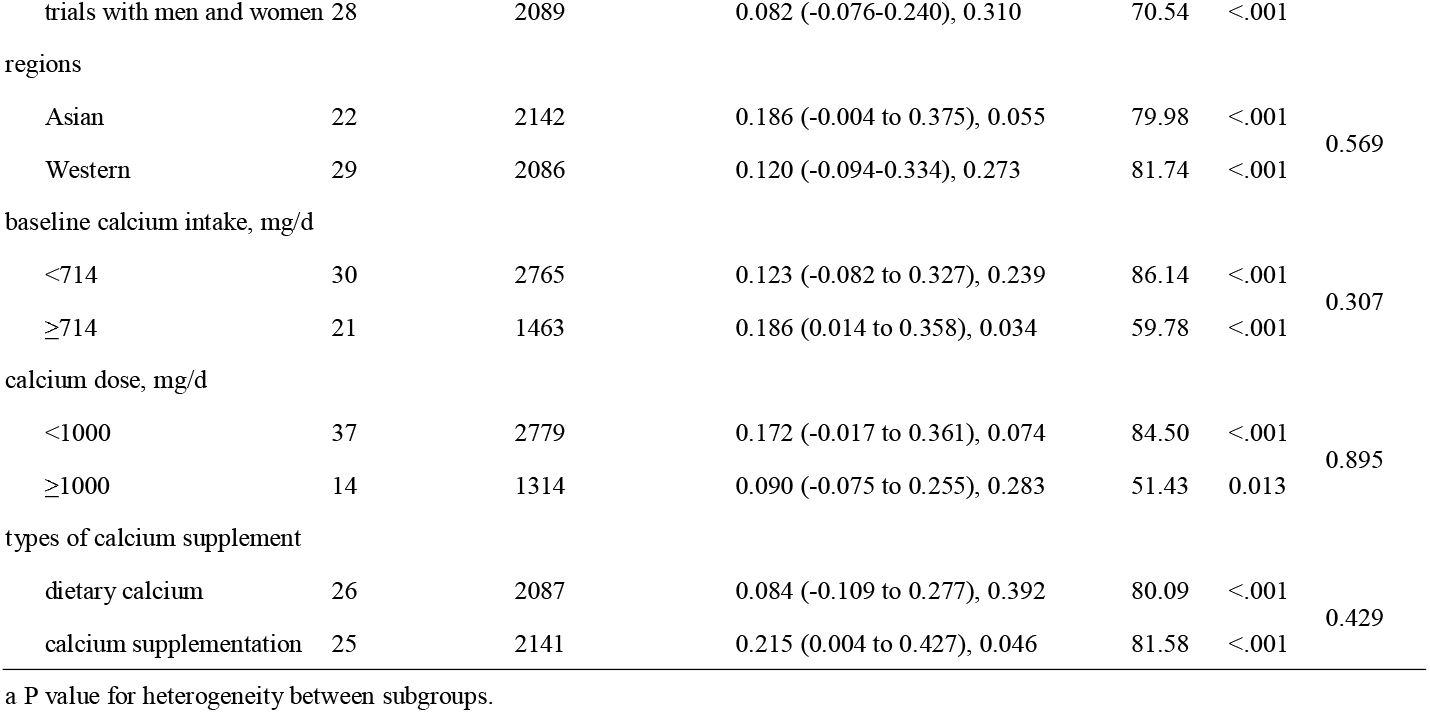
**Subgroup analysis of bone mineral content (BMC) between calcium supplementation and control for each variable at lumbar spine, femoral neck, total hip and total body**

| Variable | No. of datasets | No. of participants | BMD difference (95% CI), P Value | Heterogeneity |  | P Value<sup>a</sup> |
| --- | --- | --- | --- | --- | --- | --- |
|  |  |  |  | between studies |  |  |
|  |  |  |  | I<sup>2</sup> (%) | P-value |  |
| Lumbar Spine |  |  |  |  |  |  |
| age |  |  |  |  |  |  |
| pre-peak | 33 | 2465 | 0.173 (0.006 to 0.341), 0.043 | 75.06 | <.001 | 0.678 |
| peri-peak | 3 | 321 | 0.047 (-0.291 to 0.384), 0.786 | 47.68 | 0.148 |  |
| duration |  |  |  |  |  |  |
| <18 months | 21 | 1485 | 0.063 (-0.063 to 0.190), 0.328 | 25.21 | 0.143 | 0.487 |
| ≥18 months | 15 | 1296 | 0.293 (-0.015 to 0.602), 0.062 | 82.27 | <.001 |  |
| sex |  |  |  |  |  |  |
| women-only trials | 14 | 1220 | 0.327 (-0.017 to 0.672), 0.062 | 86.55 | <.001 | 0.496 |
| trials with men and women | 22 | 1566 | 0.076 (-0.054 to 0.207), 0.251 | 38.52 | 0.035 |  |
| regions |  |  |  |  |  |  |
| Asian | 15 | 1260 | 0.003 (-0.108 to 0.113), 0.962 | 0.00 | 0.704 | 0.112 |
| Western | 21 | 1199 | 0.319 (0.059 to 0.579), 0.016 | 82.06 | <.001 |  |
| baseline calcium intake, mg/d |  |  |  |  |  |  |
| <714 | 24 | 2030 | 0.137 (-0.075 to 0.349), 0.206 | 81.04 | <.001 | 0.104 |
| ≥714 | 12 | 756 | 0.206 (0.059 to 0.354), 0.006 | 0.00 | 0.472 |  |
| calcium dose, mg/d |  |  |  |  |  |  |
| <1000 | 29 | 2048 | 0.187 (-0.013 to 0.386), 0.067 | 78.79 | <.001 | 0.938 |
| ≥1000 | 7 | 768 | 0.097 (-0.051 to 0.245), 0.198 | 0.00 | 0.992 |  |
| types of calcium supplement |  |  |  |  |  |  |
| dietary calcium | 17 | 1267 | 0.198 (-0.119 to 0.516), 0.221 | 86.46 | <.001 | 0.447 |
| calcium supplementation | 19 | 1519 | 0.129 (0.024 to 0.234), 0.016 | 0.00 | 0.664 |  |
| Femoral Neck |  |  |  |  |  |  |
| age |  |  |  |  |  |  |
| pre-peak | 13 | 1018 | 0.249 (0.043 to 0.454), 0.018 | 58.27 | 0.004 | <.001 |
| peri-peak | 2 | 200 | 1.045 (0.701 to 1.390), <.001 | 0.00 | 0.348 |  |
| duration |  |  |  |  |  |  |
| <18 months | 9 | 648 | 0.569 (0.223 to 0.914), 0.001 | 75.38 | <.001 | 0.194 |
| ≥18 months | 6 | 570 | 0.107 (-0.062 to 0.276), 0.213 | 0.00 | 0.467 |  |
| sex |  |  |  |  |  |  |
| women-only trials | 5 | 397 | 0.742 (0.267 to 1.217), 0.002 | 74.47 | 0.004 | 0.129 |
| trials with men and women | 10 | 793 | 0.195 (-0.027 to 0.418), 0.086 | 57.60 | 0.012 |  |
| regions |  |  |  |  |  |  |
| Asian | 10 | 793 | 0.195 (-0.027 to 0.418), 0.086 | 57.60 | 0.012 | 0.129 |
| Western | 5 | 397 | 0.742 (0.267 to 1.217), 0.002 | 74.47 | 0.004 |  |
| types of calcium supplement |  |  |  |  |  |  |
| dietary calcium | 9 | 684 | 0.218 (-0.029 to 0.464), 0.083 | 60.89 | 0.009 | 0.367 |
| calcium supplementation | 6 | 506 | 0.609 (0.162 to 1.056), 0.008 | 78.02 | 0.000 |  |
| Total Hip |  |  |  |  |  |  |
| age |  |  |  |  |  |  |
| pre-peak | 13 | 1194 | 0.273 (-0.150 to 0.696), 0.206 | 91.78 | <.001 | <.001 |
| peri-peak | 1 | 121 | -1.936 (-2.346 to -1.525), <.001 | 0.00 | 1.000 |  |
| duration |  |  |  |  |  |  |
| <18 months | 6 | 542 | -0.226 (-0.514 to 0.061), 0.123 | 61.79 | 0.023 | 0.083 |
| ≥18 months | 8 | 773 | 0.385 (-0.495 to 1.264), 0.392 | 96.76 | <.001 |  |
| sex |  |  |  |  |  |  |
| women-only trials | 3 | 420 | -0.202 (-1.851 to 1.448, 0.81) | 98.13 | 0.000 | 0.499 |
| trials with men and women | 11 | 866 | 0.205 (-0.276 to 0.685), 0.404 | 91.70 | 0.000 |  |
| regions |  |  |  |  |  |  |
| Asian | 10 | 894 | 0.043 (-0.087 to 0.172), 0.516 | 0.00 | 0.691 | 0.914 |
| Western | 4 | 392 | 0.325 (-1.788 to 2.438), 0.763 | 98.71 | 0.000 |  |
| Total Body |  |  |  |  |  |  |
| age |  |  |  |  |  |  |
| pre-peak | 50 | 3762 | 0.168 (0.029 to 0.308), 0.018 | 79.47 | <.001 | <.001 |
| peri-peak | 1 | 121 | -0.716 (-1.086 to -0.347), <.001 | 0.00 | 1.000 |  |
| duration |  |  |  |  |  |  |
| <18 months | 26 | 1760 | 0.146 (-0.095 to 0.387), 0.235 | 83.36 | <.001 | 0.902 |
| ≥18 months | 25 | 2634 | 0.143 (-0.027 to 0.313), 0.100 | 77.82 | <.001 |  |
| sex |  |  |  |  |  |  |
| women-only trials | 23 | 2139 | 0.227 (-0.021 to 0.476), 0.073 | 86.47 | <.001 | 0.593 |
| trials with men and women | 28 | 2089 | 0.082 (-0.076-0.240), 0.310 | 70.54 | <.001 |  |
| regions |  |  |  |  |  |  |
| Asian | 22 | 2142 | 0.186 (-0.004 to 0.375), 0.055 | 79.98 | <.001 | 0.569 |
| Western | 29 | 2086 | 0.120 (-0.094-0.334), 0.273 | 81.74 | <.001 |  |
| baseline calcium intake, mg/d |  |  |  |  |  |  |
| <714 | 30 | 2765 | 0.123 (-0.082 to 0.327), 0.239 | 86.14 | <.001 | 0.307 |
| ≥714 | 21 | 1463 | 0.186 (0.014 to 0.358), 0.034 | 59.78 | <.001 |  |
| calcium dose, mg/d |  |  |  |  |  |  |
| <1000 | 37 | 2779 | 0.172 (-0.017 to 0.361), 0.074 | 84.50 | <.001 | 0.895 |
| ≥1000 | 14 | 1314 | 0.090 (-0.075 to 0.255), 0.283 | 51.43 | 0.013 |  |
| types of calcium supplement |  |  |  |  |  |  |
| dietary calcium | 26 | 2087 | 0.084 (-0.109 to 0.277), 0.392 | 80.09 | <.001 | 0.429 |
| calcium supplementation | 25 | 2141 | 0.215 (0.004 to 0.427), 0.046 | 81.58 | <.001 |  |

a P value for heterogeneity between subgroups.

### Publication bias

Funnel plots, Begg’s rank correlation and Egger’s regression test for each outcome bias are presented in **Supplementary file 10**. Publication bias was obvious in the femoral neck BMD. The adjusted effect size analysed using the trim and fill method also showed a difference from the unadjusted value. Except for the outcome above, no evidence for publication bias was found. The adjusted summary effect size analysed using the trim and fill method did not show substantial changes as well, which also implies no evidence of publication bias.

## Discussion

This meta-analysis comprehensively summarized the evidence for the efficiency of calcium supplementation in young people before the peak of bone mass and at the plateau period. The findings indicated significant improvement effects of calcium supplements on both BMD and BMC, especially on the femoral neck. Due to the special anatomy of the femoral neck, rotation injuries are prone to occur after partial force is applied. It is more difficult for femoral neck fractures than it is for ordinary fractures to heal after injury because the blood supply of the femoral neck is relatively poor. Overall, hip fracture is a common presentation in elderly patients and yields a 30-day mortality of approximately 8-10%.^61^ In addition, hip fracture patients are hard to care for and present a significant financial health care burden to society.^62^ Furthermore, femoral neck fracture is the most common hip fracture, accounting for 54% of hip fractures.^63^ Increasing calcium intake is likely to improve bone mass at the femoral neck, and consequently, this effect may translate into clinically meaningful reductions in hip fractures.

Numerous recent systematic reviews have concluded that there is no evidence for associations between calcium supplements and reduced risk of fracture or improvement of bone density in people aged over 50 years.^5-7 64^ Since calcium supplements are unlikely to translate into clinically meaningful reductions in fractures or improvement of bone mass in aged people, we wondered if it is possible to increase bone mass at the peak by administering calcium supplements before the age of reaching the PBM or at the plateau of this peak to prevent osteoporosis and reduce the risk of fractures in later life. To the best of our knowledge, this is the first meta-analysis to focus on age before achieving PBM or age at the plateau of PBM, at which the risk of fracture is extremely low. Why did we do such a meta-analysis? Instead of traditionally solving problems when they occurred, that is, treating osteoporosis after a patient has developed osteoporosis, our research attempted to explore the effects of preventive intervention before reaching the plateau and before osteoporosis development. Our study suggests that calcium supplementation can significantly boost peak bone content, which can improve bone mass. Since calcium supplementation in elderly individuals occurs late and has no influence, our findings have critical implications for the early prevention of fractures in the elderly population and provide better insights for the current situation of calcium supplementation. Preventive calcium supplementation in young populations is a shift in the window of intervention for osteoporosis, not limited to a certain age group but involving the whole life cycle of bone health.

Is there any difference in supplementation of calcium before or after the achievement of the PBM? We found that calcium supplementation improved the bone mass at the femoral neck in both the prepeak and peripeak subjects; furthermore, it is worth noting that the improvement effect was obviously stronger in the peripeak population (≥20-35 years) than in the prepeak population (<20 years). Based on our findings and the negative associations of calcium supplements with bone outcomes in aged people from previous studies, one can conclude that young adulthood may be the best intervention window to optimize bone mass, especially the PBM; moreover, our study indicates the importance of calcium supplementation at this age instead of the often-mentioned age groups of children or elderly individuals. The findings of our study provide completely new insight into a novel intervention window in young adulthood to improve bone mass and further prevent osteoporosis and fractures in their late lifespan. To synthesize previously published studies in children, we found a meta-analysis conducted by Winzenberg et al^13^ that included 19 studies involving 2859 children and found a small effect on total body BMC and no effect on lumbar spine BMD in children, which was in line with our finding. However, they found no effect on BMD at the femoral neck, which was inconsistent with our result. We therefore performed a sensitivity analysis, excluding all the literature they included, and found that the results of our newly included studies, 28 in total, were generally consistent with the primary results. We also performed a sensitivity analysis incorporating only the studies they pooled and found a statistically significant effect for BMD in the femoral neck and total body, while the results for total body BMC were nonsignificant. (see eTable 6 and 7) These slightly different findings can be interpreted as follows: first, we included more and updated literature; second, they used only endpoint data directly, whereas we used change data, taking into account the difference in baseline conditions; third, we used change data to represent the change before and after calcium supplementation more directly. Another meta-analysis conducted by Huncharek et al^14^ included 21 studies involving 3821 subjects and pooled three reports involving subjects with low baseline calcium intake and reported a statistically significant summary of the mean BMC in children. Combining the above published literature with our conclusions, it can be concluded that calcium supplementation is more effective in young adults aged 20-35 years than in children. Although this issue needs to be confirmed in the future, our findings highlight the importance of this intervention window of approximately 10-15 years at the peri-PBM period, which is better than the pre-PBM period.

To explore whether there is a difference between dietary calcium intake and calcium supplements, our subgroup analyses suggested that one can obtain this beneficial effect from both calcium sources, including dietary intake and calcium supplements. For BMD at the femoral neck, dietary calcium seemed to exert a better effect than calcium supplements. Similarly, we also found that the improvement effect was statistically significant only in subjects supplied with calcium dosages lower than 1000 mg/d. These findings support the hypothesis that there may be a threshold dose of calcium supplementation; when exceeded, the effect does not increase. Our findings are consistent with the previous research by Prentice et al, which is that no additional benefit is associated with an intake above the currently recommended dose at the population level.^65^ The underlying mechanisms are unclear and need to be elucidated in future studies.

To determine the differences between high dietary intake and low dietary intake of calcium at baseline, our subgroup analyses showed that the improvement effect seemed to be stronger in subjects with high intake at baseline than in those in the lower subgroup. Interestingly, these results were in accordance with the findings of subgroup analyses by population area, which suggested that calcium supplementation was more effective in Western populations, whose level of baseline calcium intake is normally higher than that in Asian countries. However, these findings are likely to be contrary to our common sense, which is, that under normal circumstances, the effects of calcium supplementation should be more obvious in people with lower calcium intake than in those with higher calcium intake. Therefore, this issue needs to be tested and confirmed in future trials.

To investigate changes in the effect of calcium supplementation after cessation, our subgroup analysis showed that the effect remained significant 1 year after cessation, particularly at various sites of BMD. For studies with a follow-up period longer than one year, we included only two articles: one study^53^ with two years of follow-up after calcium supplementation was stopped and another study^58^ with seven years of follow-up. Their results were pooled and showed that the effects of calcium supplementation no longer persisted. The number of studies is too small for us to explore how long the effects of calcium supplementation will last, and well-designed cohort studies are needed in the future. In the meantime, we have found a point to ponder about whether gains can be made when calcium supplementation is restarted after a period of withdrawal and what other changes in the organism remain to be discovered.

Several limitations need to be considered. First, there was substantial intertrial heterogeneity in the present analysis, which might be attributed to the differences in baseline calcium intake levels, regions and sexes according to subgroup and meta-regression analyses. To take heterogeneity into account, we used random effect models to summarize the effect estimates, which could reduce the impact of heterogeneity on the results to some extent. Second, our research failed to clearly compare the difference between males and females due to the limitation of existing data-- some studies provided merged data of males and females without males alone. Based on the existing data, the beneficial effect was more obvious when subjects were limited to women only, which needs to be validated in future trials. Third, we found that few of the existing studies focused on the 20-35-year age group, which was why there were only three studies of this age group that met our inclusion criteria; although the number was small, our evidence was of high quality, and the results were stable, especially in the femoral neck. We also tried to find mechanisms related to bone metabolism in the age group of 20-35 years, but few studies have focused on this age group; most studies have focused only on mechanisms related to older people or children. Therefore, more high-quality RCTs and studies on the exploration of mechanisms focusing on the 20-35-year age group are needed in the future. Finally, as some of the studies did not provide the physical activity levels of the participants, we failed to exclude the effect of physical activity on the results.

This study has several strengths. In this first systematic review by meta-analysis to focus on people at the age before achieving PBM and at the age around the peak of bone mass, we comprehensively searched for all of the currently eligible trials and included a total of 7382 participants (including 3283 calcium supplement users and 4099 controls), which added reliability to our findings. Another strength is the high consistency of the results across predesigned subgroup analyses and sensitivity analyses. Additionally, we analysed both BMD and BMC separately for the different measurement sites rather than using the mean of all combined values to draw conclusions, which has the advantage of obtaining changes in bone indexes at different sites and drawing more accurate conclusions.

In conclusion, calcium supplementation can significantly improve BMD and BMC, especially at the femoral neck. Moreover, supplementation in people who are at the plateau of their PBM has a better effect. Although further well-designed RCTs with larger sample sizes are required to verify our findings, we provide a new train of thought regarding calcium supplementation and the evaluation of its effects. In terms of bone health and the full life cycle of a person, the intervention window of calcium supplementation should be advanced to the age around the plateau of PBM, namely, at 20-35 years of age.

## Data Availability

All data in this analysis are based on published studies. Supplementary data files contain all raw tabulated data are provided in appendix.

## Contributors

SRW had full access to all the data in the study and takes responsibility for the integrity of the data and the accuracy of the data analysis. SRW and YPL developed the initial idea for the study. SYL and YPL organised the study concept and design. YPL, SYL and YL acquired, analysed, and interpreted the data. YPL and SYL did statistical analysis. YPL, SYL, YL, HNJ, BYR, YFH, XMA, XDS, XYF and SRW drafted the manuscript.

## Acknowledgments

We thank all authors and participants of the original studies. We also thank American Journal Experts for English language polishing of the manuscript.

## What is already known on this topic

- A number of meta-analyses have demonstrated that calcium supplementation is not associated with lower fracture incidence or bone mineral accretion. The results of studies on the effectiveness of calcium supplementation in young populations were inconsistent. Most studies focused on children or adolescent, rather than young adults under 35 years old-- from the perspective of peak bone mass (PBM). Although each of these studies has contributed imperative findings to the field, the effect of calcium supplementation in people at the age before or around achieving PBM has remained unanswered.

## What are the findings

- Overall, moderate certainty evidence suggests that calcium supplementation can significantly improve the BMD levels of total body and femoral neck, slightly increase the BMC level of femoral neck, total body and lumbar spine. The findings were robust in extensive pre-specific sensitivity analyses and subgroup analyses. Additionally, the improvement in bone mass of femoral neck was more pronounced in the peri-PBM population (20-35 years) than the pre-PBM population (<20 years).

## How might it impact on clinical practice in the near future

- This study is a comprehensive and robust original evidence-based study that focused on calcium supplementation in young populations, which is important to an international general medical audience, to public health, or to policy decisions worldwide.
- Over the past decades, the intervention window for osteoporosis focused on the elderly population, however, in recent years, several studies have concluded that calcium supplementation in the elderly does not prevent osteoporosis or reduce risk of fracture. Our findings provided novel insights and evidence in calcium supplementation, which showed that calcium supplementation significantly improves bone mass, implying that preventive calcium supplementation before or around achieving PBM may be a shift in the window of intervention for osteoporosis.

---

**Figure 2-figure supplement 1A.**
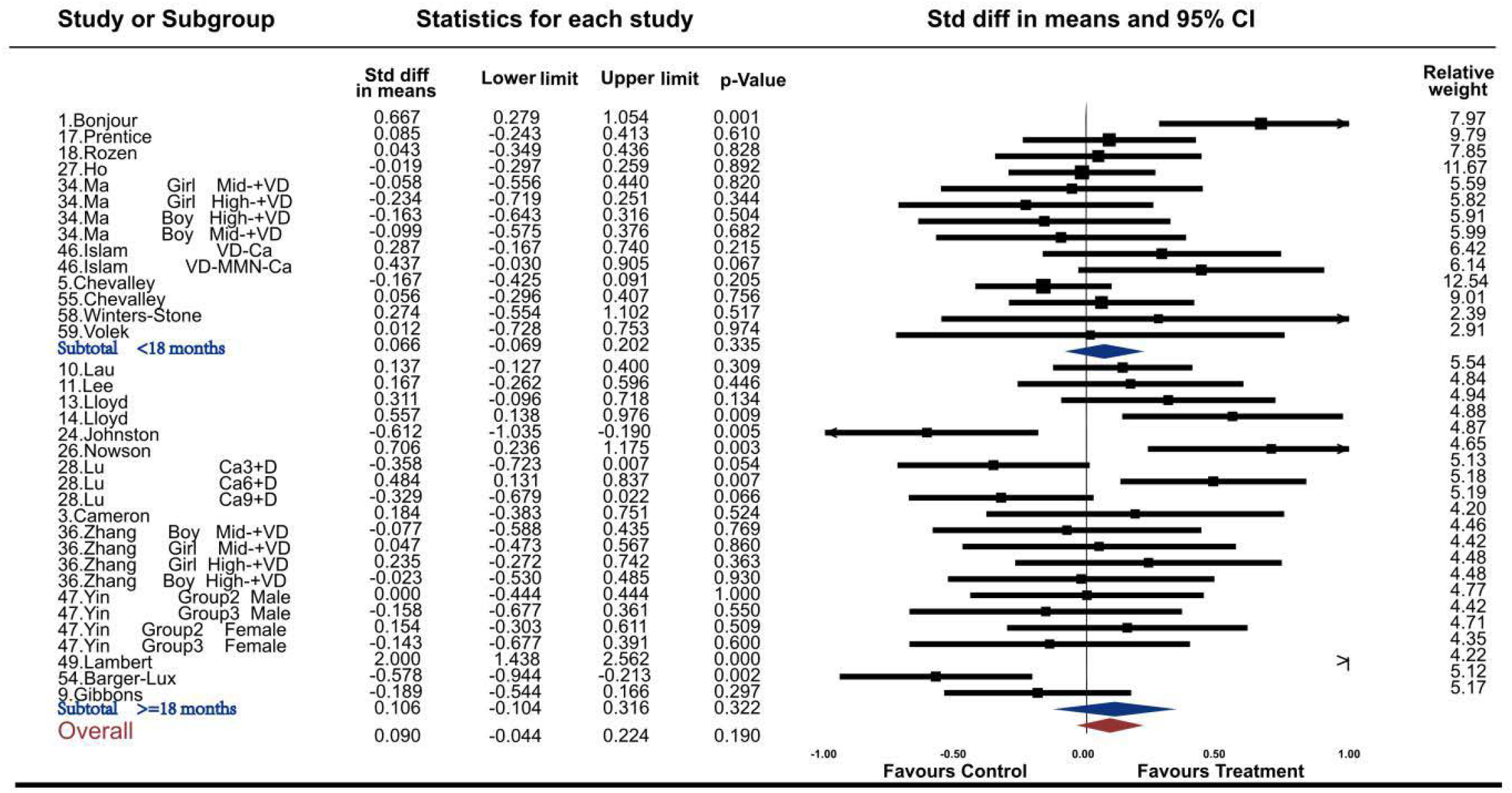
Forest plots for the association between calcium supplementation and the accretion of lumbar spine bone mineral density (LSBMD)

**Figure 2-figure supplement 1B.**
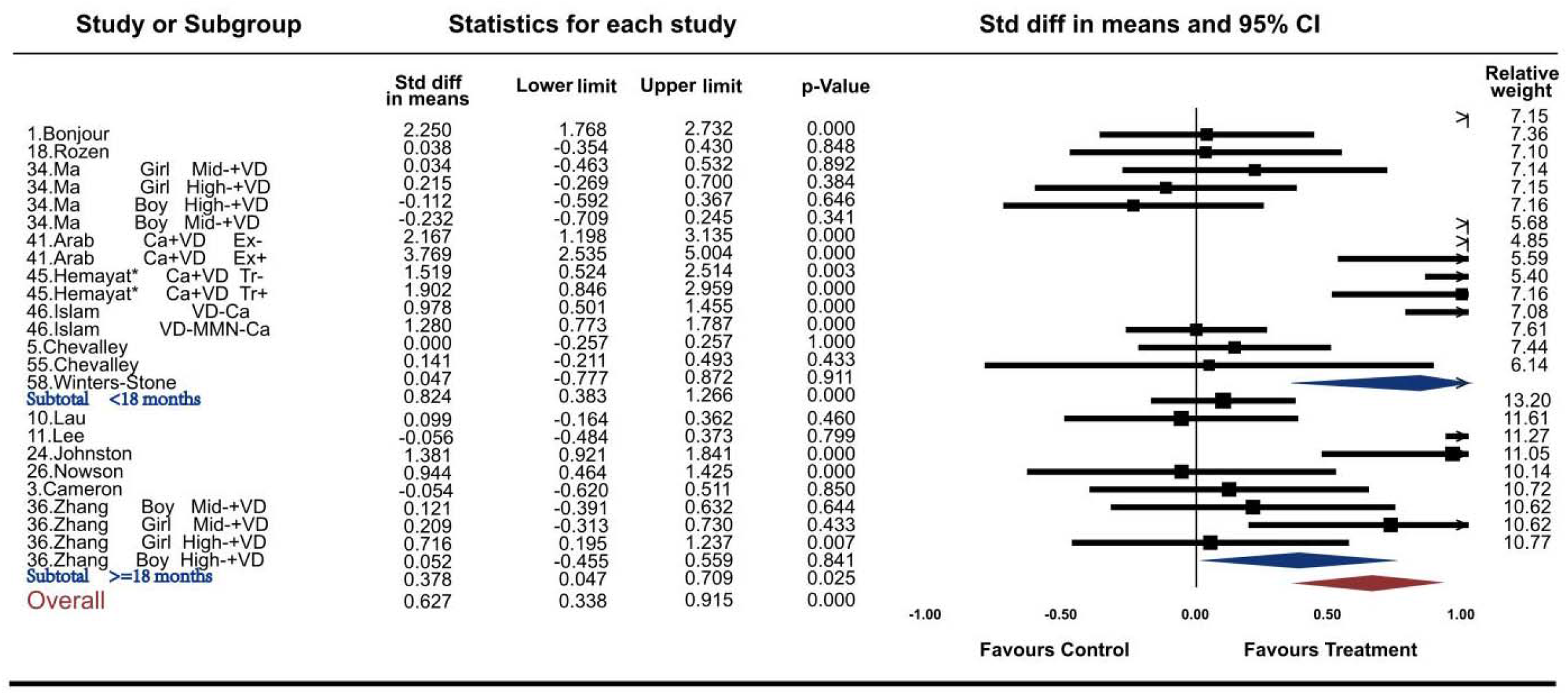
Forest plots for the association between calcium supplementation and the accretion of femoral neck bone mineral density (FNBMD)

**Figure 2-figure supplement 1C.**
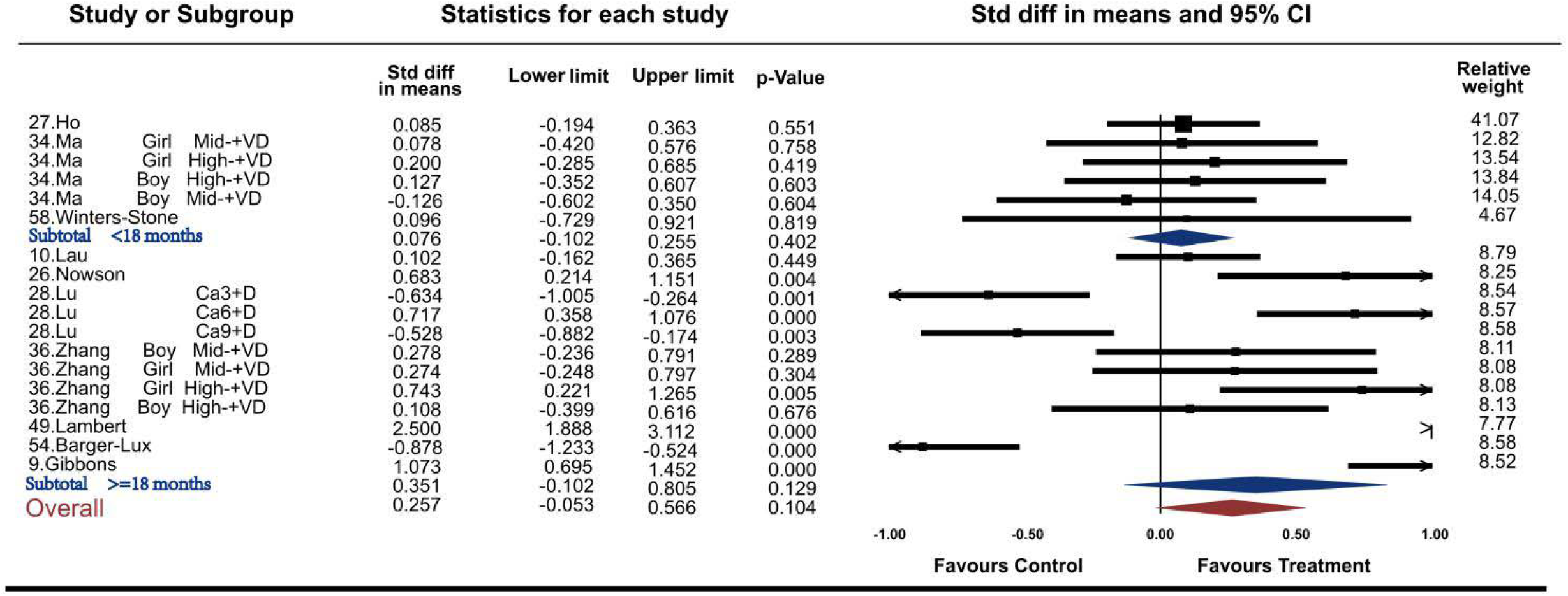
Forest plots for the association between calcium supplementation and the accretion of total hip bone mineral density (THBMD)

**Figure 2-figure supplement 1D.**
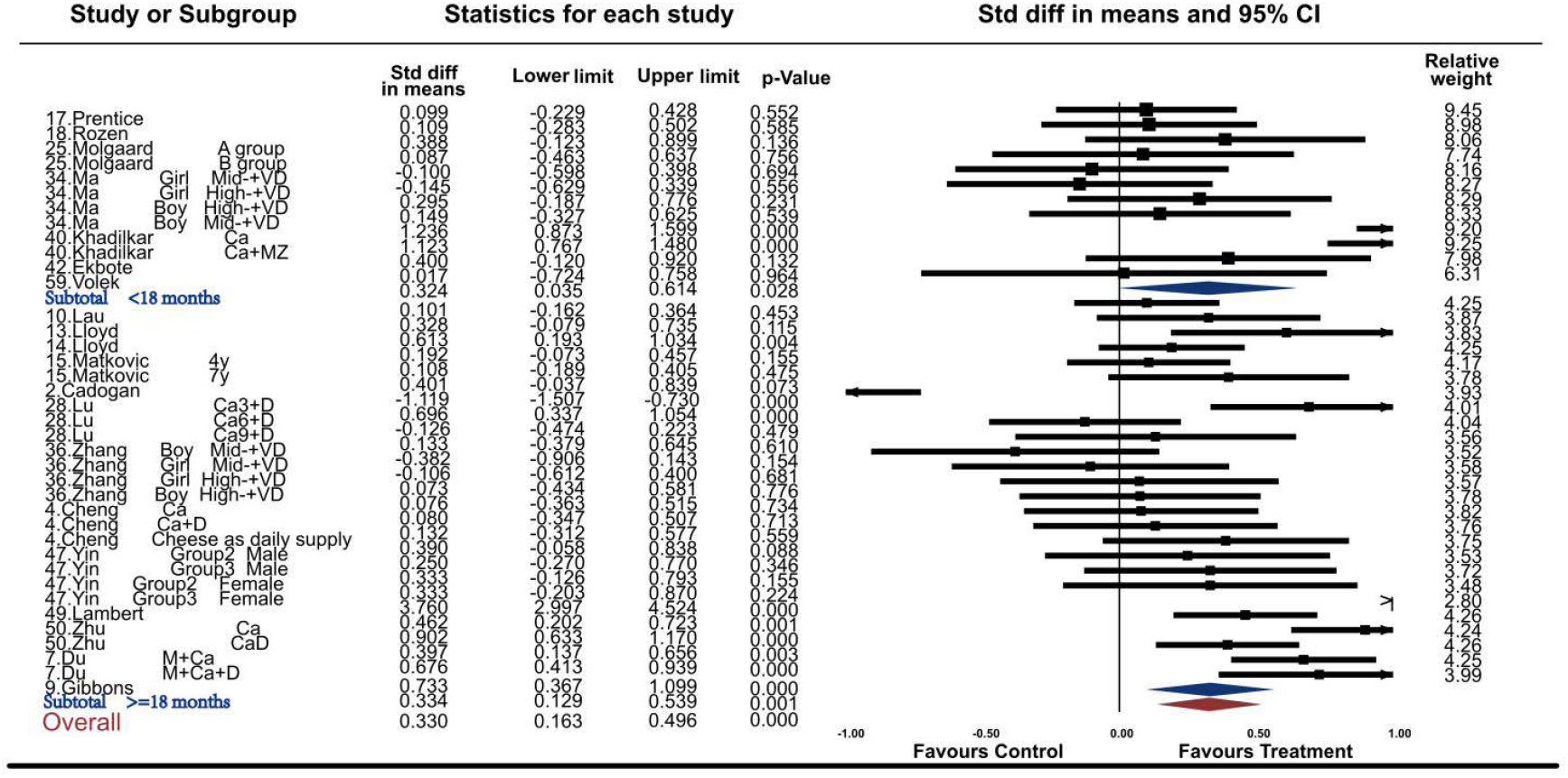
Forest plots for the association between calcium supplementation and the accretion of total body bone mineral density (TBBMD)

**Figure 3-figure supplement 1A.**
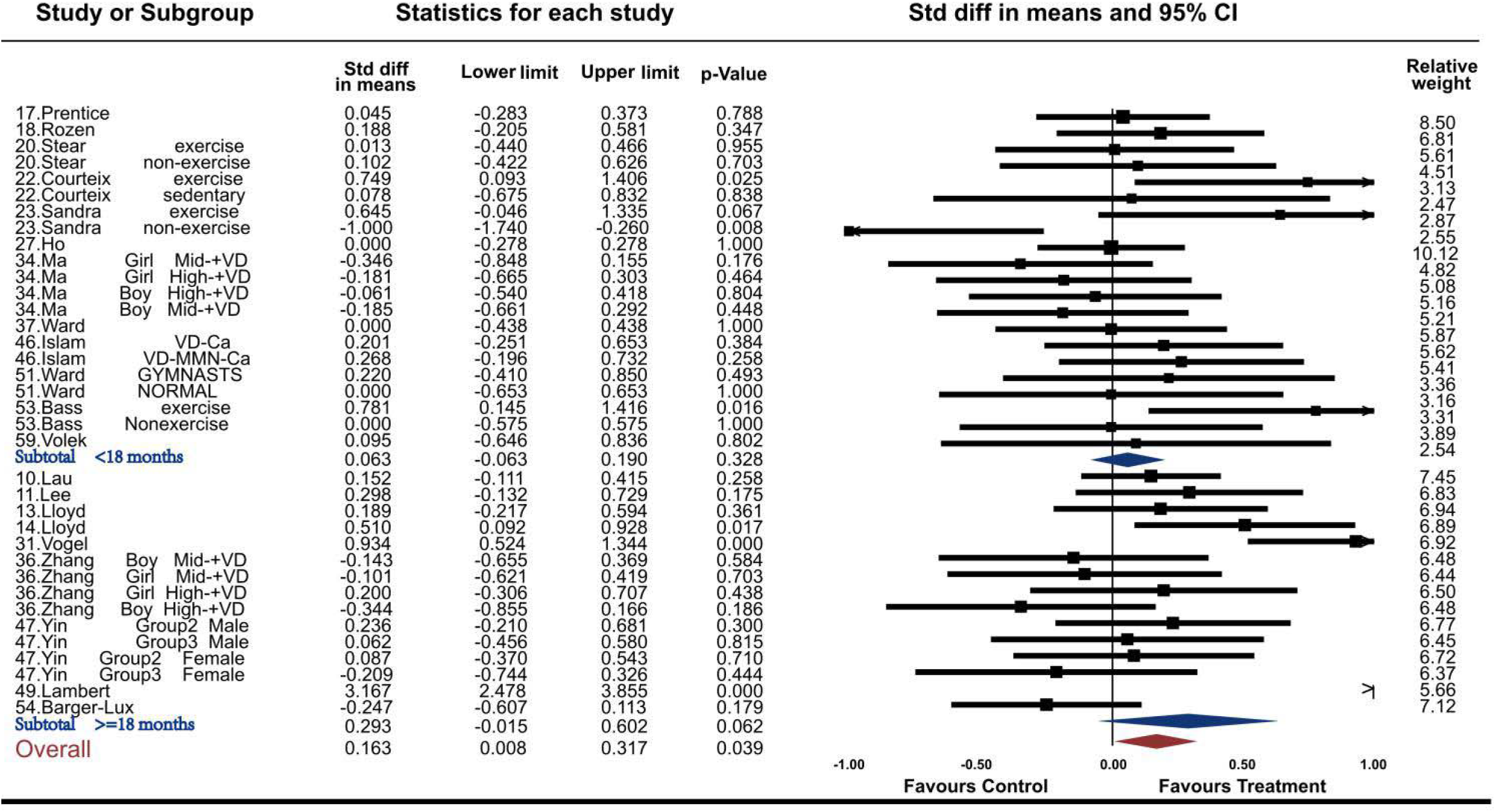
Forest plots for the association between calcium supplementation and the accretion of lumbar spine bone mineral content (LSBMC)

**Figure 3-figure supplement 1B.**
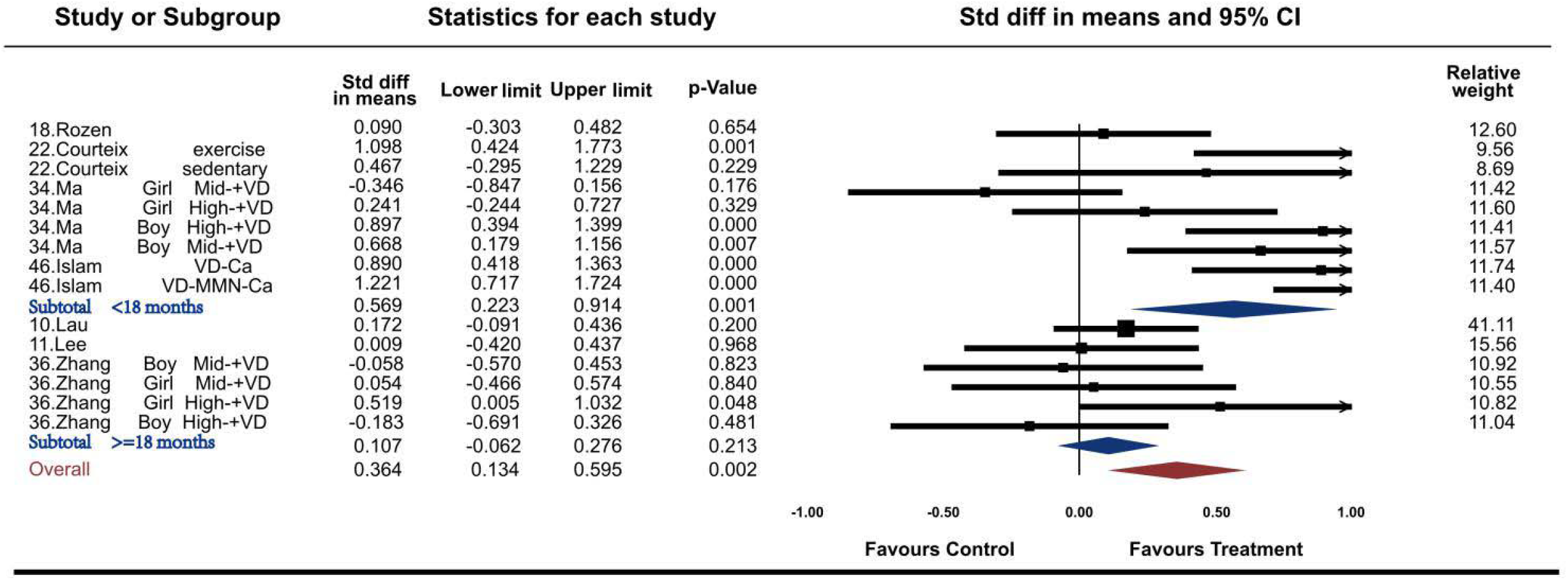
Forest plots for the association between calcium supplementation and the accretion of femoral neck bone mineral content (FNBMC)

**Figure 3-figure supplement 1C.**
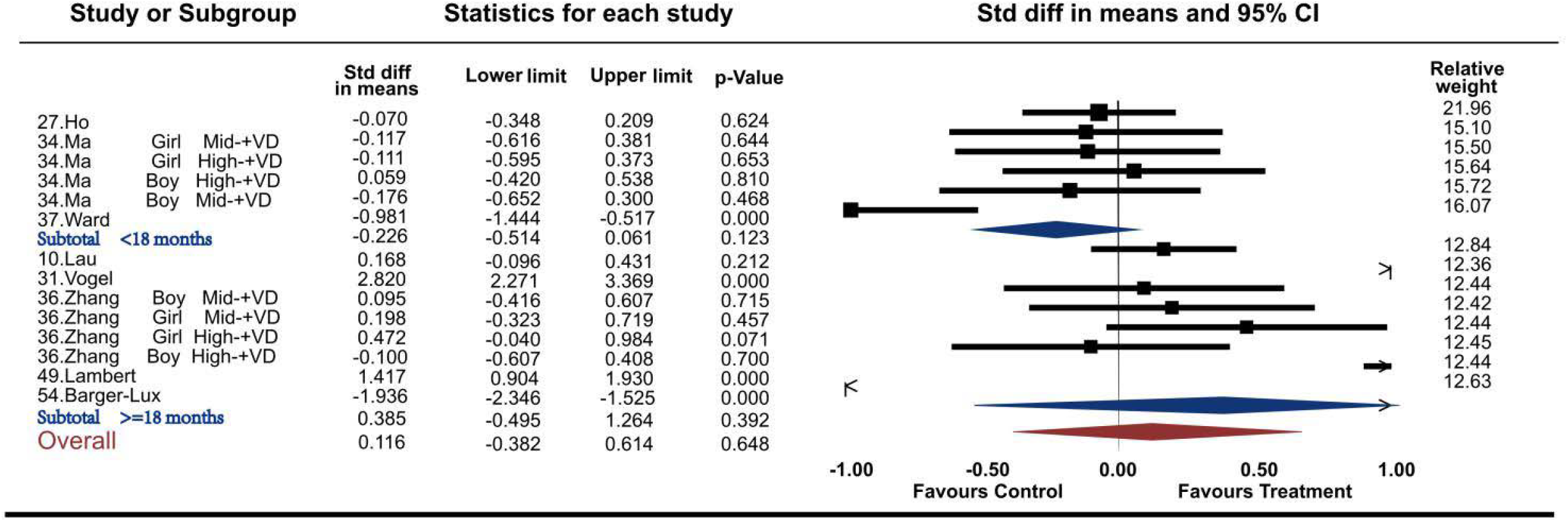
Forest plots for the association between calcium supplementation and the accretion of total hip bone mineral content (THBMC)

**Figure 3-figure supplement 1D.**
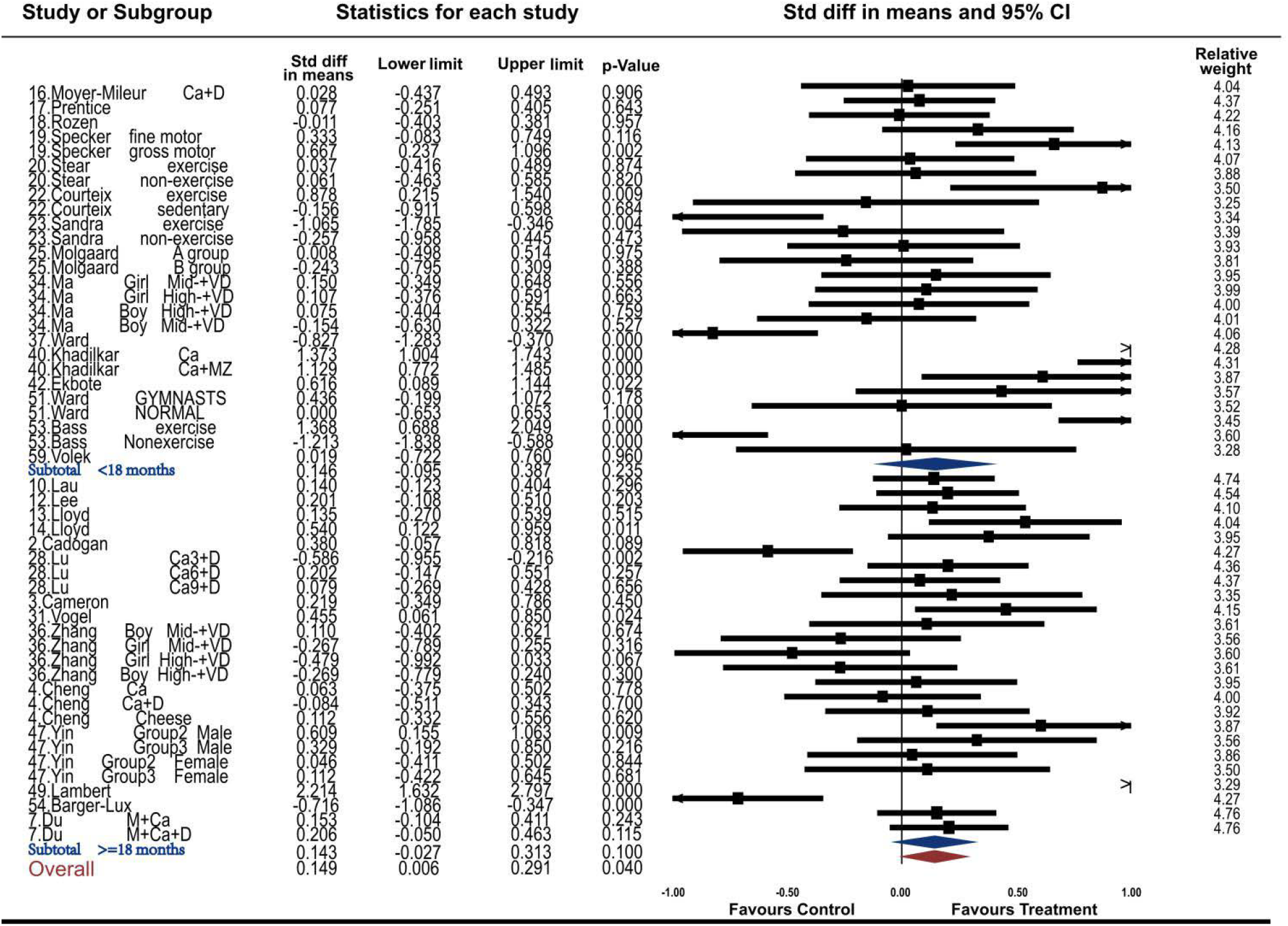
Forest plots for the association between calcium supplementation and the accretion of total body bone mineral content (TBBMC)

