## Supplementary file 1 for "The effect of calcium supplementation in people under 35 years old: A systematic review and meta-analysis of randomized controlled trials"

**Supplementary file 1**: Search strategies

*Pubmed*

#1. **(((((((((((calcium[MeSH Terms]) OR (calcium[Title/Abstract])) OR (calcium carbonate[Title/Abstract])) OR (calcium citrate[Title/Abstract])) OR (calcium pills[Title/Abstract])) OR (calcium supplement[Title/Abstract])) OR (Ca2[Title/Abstract])) OR (dairy product[Title/Abstract])) OR (milk[Title/Abstract])) OR (yogurt[Title/Abstract])) OR (cheese[Title/Abstract])) OR (dietary supplement[Title/Abstract])**

#2. **(((randomized controlled trial[Publication Type]) OR (randomized controlled trial[Title/Abstract])) OR (clinical trials[Title/Abstract])) OR (RCT[Title/Abstract])**

#3. #1 AND #2

#4. **(((((Bone Density[MeSH Terms]) OR (bone density[Title/Abstract])) OR (bone mineral density[Title/Abstract])) OR (bone mineral densities[Title/Abstract])) OR (bone mineral content[Title/Abstract])) OR (bone mineral contents[Title/Abstract])**

#5. #3 AND #4

*EMBASE*

#1. ’**calcium’**:ti,ab,kw OR *’***calcium carbonate’**:ti,ab,kw OR ’**calcium citrate’**:ti,ab,kw OR ’**calcium pills’**:ti,ab,kw OR *’***calcium supplement’**:ti,ab,kw OR *’***Ca2’**:ti,ab,kw OR *’***dairy product’**:ti,ab,kw OR *’***milk’**:ti,ab,kw OR *’***yogurt’**:ti,ab,kw OR *’***cheese’**:ti,ab,kw OR *’***dietary supplement’**:ti,ab,kw

#2. *’***randomized controlled trial’**:ti,ab,kw OR *’***clinical trials’**:ti,ab,kw OR ’ RCT **’**:ti,ab,kw

#3. #1 AND #2

#4. ’**bone density’**:ti,ab,kw OR *’***bone mineral density’**:ti,ab,kw OR *’***bone mineral densities’**:ti,ab,kw OR *’***bone mineral content’**:ti,ab,kw OR *’***bone mineral contents’**:ti,ab,kw

#5. #3 AND #4

*ProQuest*

#1. mesh(**calcium) OR mainsubject(calcium) OR ab(calcium carbonate) OR ab(calcium citrate) OR ab(calcium pills) OR ab(calcium supplement) OR ab(Ca2) OR ab(dairy product) OR ab(milk) OR ab(yogurt) OR ab(cheese) OR ab(dietary supplement)**

**#2. mesh(Randomized Controlled Trials as Topic)** OR mesh(controlled clinical trials as topic) OR ab(Randomized Controlled Trial) OR ab(controlled clinical trial) OR ab(controlled trial) OR ab(clinical trial) OR ab(RCT)

#3. #1 AND #2

**#4 mesh(Bone Density) OR mainsubject(bone density) OR ab(bone mineral density) OR ab(bone mineral densities) OR ab(bone mineral content) OR ab(bone mineral contents)**

#5. #3 AND #4

*CENTRAL (Cochrane Central Register of Controlled Trials)*

#1. MeSH descriptor: [calcium] explode all trees

#2. (**calcium carbonate)**:ti,ab,kw OR (**calcium citrate)**:ti,ab,kw OR (**calcium pills)**:ti,ab,kw OR (**calcium supplement)**:ti,ab,kw OR (**Ca2)**:ti,ab,kw OR (**dairy product)**:ti,ab,kw OR (**milk)**:ti,ab,kw OR (**yogurt)**:ti,ab,kw OR (**cheese)**:ti,ab,kw OR (**dietary supplement)**:ti,ab,kw

#3. #1 OR #2

#4. MeSH descriptor: [**Bone Density] explode all trees**

**#5. (bone density)**:ti,ab,kw OR (**bone mineral density)**:ti,ab,kw OR (**bone mineral densities)**:ti,ab,kw OR (**one mineral content)**:ti,ab,kw OR (**bone mineral contents)**:ti,ab,kw

#6. #4 OR #5

#7. #3 AND #6
