## Supplementary file 2 for "The effect of calcium supplementation in people under 35 years old: A systematic review and meta-analysis of randomized controlled trials"

| **Supplementary file 2. Excluded Trials and Reasons for Exclusion** | |
| --- | --- |
| **Excluded trials** | **Reasons for exclusion** |
| Grados 2003 | This randomised trial enrolled participants aged over 65 years old. |
| Chee 2003 | This randomised trial enrolled participants in postmenopausal. |
| Meier 2004 | This randomised trial enrolled participants aged around 60 years old. |
| Pongchaiyakul 2004 | This is a cross-sectional study and participants aged over 35 years old. |
| Daniele 2004 | This randomised trial enrolled participants in peri- and post-menopause. |
| Daly 2006 | This randomised trial enrolled older men. |
| Jackson 2006 | This randomised trial enrolled participants in postmenopausal. |
| Prince 2006 | This randomised trial enrolled elderly women. |
| Daly 2006 | This randomised trial enrolled older men. |
| Reid 2006 | This randomised trial enrolled older men. |
| Moschonis 2006 | This randomised trial enrolled participants in postmenopausal. |
| Bolton-Smith 2007 | This randomised trial enrolled older women. |
| Zhu 2007 | This randomised trial enrolled elderly women. |
| Kukuljan 2009 | This randomised trial enrolled older men. |
| Reid 2008 | This randomised trial enrolled older men. |
| Vujasinović-Stupar 2009 | This randomised trial enrolled participants in postmenopausal. |
| Kärkkäinen 2010 | This randomised trial enrolled women aged 65-71 years. |
| Thomas 2010 | This randomised trial enrolled overweight premenopausal women. |
| Tenta 2010 | This randomised trial enrolled participants in postmenopausal. |
| Jackson 2011 | This randomised trial enrolled participants in older postmenopausal women. |
| Moschonis 2011 | This randomised trial enrolled participants in older postmenopausal women. |
| Gui 2012 | This randomised trial enrolled participants in older postmenopausal women. |
| Nakamura 2012 | This randomised trial enrolled participants in perimenopausal and postmenopausal Asian women. |
| Rajatanavin 2013 | This randomised trial enrolled elderly women. |
| Radford 2014 | This randomised trial enrolled participants in older postmenopausal women. |
| Kruger 2016 | This randomised trial enrolled participants in pre- and postmenopausal. |
| Mathis 2015 | This randomised trial enrolled participants aged over 35 years old. |
| Chen 2016 | This randomised trial enrolled participants in older postmenopausal women. |
| Sakai 2017 | This randomised trial enrolled participants in older postmenopausal women. |
| Sullivan 2017 | This randomised trial enrolled participants in older postmenopausal women. |
| Bristow 2017 | This randomised trial enrolled participants aged over 35 years old. |
| Kruger 2018 | This randomised trial enrolled participants in older postmenopausal women. |
| Reyes-Garcia 2018 | This randomised trial enrolled participants in older postmenopausal women. |
| Barnuevo 2018 | This randomised trial enrolled participants in older premenopausal women. |
| Nakamura 2019 | This randomised trial enrolled participants in peri and postmenopausal. |
| Ilich 2019 | This randomised trial enrolled participants in older premenopausal women. |
| Zhang 2020 | This randomised trial enrolled participants in perimenopausal. |
| Morato-Martínez 2020 | This randomised trial enrolled participants in high-risk menopausal. |
| Fuleihan 2006 | This study was vitamin D monotherapy without calcium. |
| Dibba 2000 | No available bone measurement data. |
| Viljakainen 2006 | This study was vitamin D monotherapy without calcium. |
| Gaffney-Stomberg 2014 | No available bone measurement data. |
| Cullers 2019 | Participants were during pregnancy. |
| Normando 2016 | Participants were during pregnancy. |
| Zhang 2016 | Participants were in the status of postpartum lactating. |
| Gaffney-Stomberg 2019 | No available bone measurement data. |
| Jarjou 2013 | Participants were during pregnancy. |
| Diogenes 2013 | Participants were during pregnancy. |
| Jarjou 2010 | Participants were during pregnancy. |
| Liu 2010 | Participants were during pregnancy. |
| Woo 2007 | No available data after application. |
| Matkovic 2004 | Repeated data sources with the publication included in the text from Matkovic et al in 2005. |
| Mehlenbeck 2004 | No available data after application. |
| Yu 2011 | This study focused on specific genotype. |
| Ekbote 2013 | This study focused mainly on growth hormone-deficient children. |
| Vecchi 2012 | This study only enrolled HIV-infected patients, and was cross-sectional study. |
| Bosworth 2012 | The participants were over 35 years old and had chronic kidney disease. |
| Arpadi 2012 | This study only enrolled HIV-infected patients. |
| Sang Hyeon 2011 | This study enrolled participants aged over 35 years old and used Vitamin K supplement along with vitamin D and calcium . |
| Min-Yu 2015 | The participants were osteoporotic patients. |
| Thacher 2016 | This study focused on nutritional rickets. |
| Pignotti 2010 | This study was vitamin D monotherapy without calcium. |
| Chailurkit 2010 | This study was vitamin D monotherapy without calcium. |
| Aloia 2010 | No available bone measurement data. |
