## Supplementary file 3 for "The effect of calcium supplementation in people under 35 years old: A systematic review and meta-analysis of randomized controlled trials"

**Supplementary file 3. Risk of bias assessment for eligible trials**

| Study ID | Weight | Randomization process | Deviations from intended interventions | Mising outcome data | Measurement of the outcome | Selection of the reported result | Overall Bias |
| --- | --- | --- | --- | --- | --- | --- | --- |
| Bonjour 1997 | 1 | Low | Low | Low | Low | Low | Low |
| Cadogan 1997 | 1 | Low | Low | Low | Low | Some concerns | Some concerns |
| Cameron 2004 | 1 | Low | Low | Low | Low | Some concerns | Some concerns |
| Cheng 2005 | 1 | Low | Low | Low | Low | Low | Low |
| Chevalley 2005 | 1 | Low | Low | Low | Low | Low | Low |
| Du 2004 | 1 | Low | Low | Low | Low | Some concerns | Some concerns |
| Gibbons 2004 | 1 | Low | Low | Low | Low | Some concerns | Some concerns |
| Lau 2004 | 1 | Low | Low | Low | Low | Low | Low |
| Lee 1995 | 1 | Low | Low | Low | Low | Low | Low |
| Lee 1994 | 1 | Low | Low | Low | Low | Low | Low |

**Supplementary file 3. Risk of bias assessment for eligible trials (continued)**

| Lloyd 1994 | 1 | Low | Low | Low | Low | Low | Low |
| --- | --- | --- | --- | --- | --- | --- | --- |
| Lloyd 1996 | 1 | Low | Low | Low | Low | Low | Low |
| Matkovic 2005 | 1 | Low | Low | Low | Low | Low | Low |
| Moyer-Mileur 2003 | 1 | Low | Low | Low | Low | Some concerns | Some concerns |
| Prentice 2005 | 1 | Low | Low | Low | Low | Low | Low |
| Rozen 2003 | 1 | Low | Low | Low | Low | Low | Low |
| Specker 2003 | 1 | Low | Low | Low | Some concerns | Low | Some concerns |
| Stear 2003 | 1 | Low | Low | Low | Low | Low | Low |
| Courteix 2005 | 1 | Low | Low | Low | Low | Low | Low |
| Sandra 2003 | 1 | Low | Low | Low | Some concerns | Low | Some concerns |
| Johnston 1992 | 1 | Low | Low | Low | Low | Low | Low |
| Molgaard 2004 | 1 | Low | Low | Low | Low | Some concerns | Some concerns |
| Nowson 1997 | 1 | Low | Low | Low | Low | Low | Low |

**Supplementary file 3. Risk of bias assessment for eligible trials (continued)**

| Ho 2005 | 1 | Low | Some concerns | Low | Low | Low | Some concerns |
| --- | --- | --- | --- | --- | --- | --- | --- |
| Lu 2019 | 1 | Low | Low | Low | Some concerns | Low | Some concerns |
| Vogel 2017 | 1 | Low | Some concerns | Low | Some concerns | Low | Some concerns |
| Ma 2014 | 1 | Low | Low | Low | Low | Low | Low |
| Zhang 2014 | 1 | Low | Low | Low | Low | Low | Low |
| Ward 2014 | 1 | Low | Low | Low | Some concerns | Low | Some concerns |
| Khadilkar 2012 | 1 | Low | Low | Low | Low | Low | Low |
| Arab 2012 | 1 | Some concerns | High | Low | Some concerns | Low | High |
| Ekbote 2011 | 1 | Some concerns | Some concerns | Low | Low | Low | Some concerns |
| Hemayattalab 2010 | 1 | Some concerns | High | Low | Some concerns | Low | High |
| Islam 2010 | 1 | Low | Low | Low | Some concerns | Low | Some concerns |
| Yin 2010 | 1 | Some concerns | Some concerns | Low | Low | Low | Some concerns |
| Lambert 2008 | 1 | Low | Low | Low | Low | Low | Low |

**Supplementary file 3. Risk of bias assessment for eligible trials (continued)**

| Zhu 2008 | 1 | Low | Low | Low | High | Some concerns | High |
| --- | --- | --- | --- | --- | --- | --- | --- |
| Ward 2007 | 1 | Low | Low | Low | Low | Low | Low |
| Bass 2007 | 1 | Low | Low | Low | Low | Low | Low |
| Barger-Lux 2005 | 1 | Low | Low | Low | Low | Low | Low |
| Chevalley 2005 | 1 | Low | Low | Low | Low | Low | Low |
| Winters-Stone 2004 | 1 | Some concerns | Low | Some concerns | Some concerns | Low | Some concerns |
| Volek 2003 | 1 | Some concerns | High | Low | Low | Low | High |
