## Supplementary file 4 for "The effect of calcium supplementation in people under 35 years old: A systematic review and meta-analysis of randomized controlled trials"

**Supplementary file 4A. Sensitivity analyses excluding studies of low or medium quality in bone mineral density (BMD)**

| **Subgroups** | **No. of studies or subgroups** | **Effect estimate (95%CI), P-value** | | **Heterogeneity between studies** | | **P-value for heterogeneity between subgroups** |
| --- | --- | --- | --- | --- | --- | --- |
|  |  |  |  | **I**² (%) | **P-value** |  |
| Total | 54 | 0.413 (0.261 to 0.565), <.001 | | 86.28 | <.001 | 0.988 |
| Total* | 28 | 0.387 (0.170 to 0.605), <.001 | | 88.13 | <.001 |  |
| **Lumbar Spine** |  |  |  |  |  |  |
| Subtotal | 35 | 0.090 (-0.044 to 0.224), 0.190 | | 71.89 | <.001 | 0.84 |
| Subtotal* | 21 | 0.13 (-0.068 to 0.327), 0.198 | | 79.52 | <.001 |  |
| **Femoral Neck** |  |  |  |  |  |  |
| Subtotal | 24 | 0.627 (0.338 to 0.915), <.001 | | 88.27 | <.001 | 0.382 |
| Subtotal* | 16 | 0.356 (0.064 to 0.648), 0.017 | | 87.09 | <.001 |  |
| **Total Hip** |  |  |  |  |  |  |
| Subtotal | 18 | 0.257 (-0.053 to 0.566), 0.104 | | 89.68 | <.001 | 0.776 |
| Subtotal* | 12 | 0.320 (-0.075 to 0.715), 0.112 | | 89.08 | <.001 |  |
| **Total Body** |  |  |  |  |  |  |
| Subtotal | 38 | 0.330 (0.163 to 0.496), <.001 | | 85.15 | <.001 | 0.944 |
| Subtotal* | 21 | 0.343 (0.098 to 0.588), 0.006 | | 86.9 | <.001 |  |

*The pooled results of sensitivity analyses excluding the studies of low or medium quality.

**Supplementary file 4B. Sensitivity analyses excluding studies of low or medium quality in bone mineral content (BMC)**

| **Subgroups** | **No. of studies or subgroups** | **Effect estimate (95%CI), P-value** | | **Heterogeneity between studies** | | **P-value for heterogeneity between subgroups** |
| --- | --- | --- | --- | --- | --- | --- |
|  |  |  |  | **I² (%)** | **P-value** |  |
| Total | 55 | 0.285 (0.154 to 0.415), <.001 | | 79.28 | <.001 | 0.749 |
| Total* | 30 | 0.354 (0.152 to 0.556), 0.001 | | 83.32 | <.001 |  |
| **Lumbar Spine** |  |  |  |  |  |  |
| Subtotal | 36 | 0.163 (0.008 to 0.317), 0.039 | | 73.71 | <.001 | 0.977 |
| Subtotal* | 24 | 0.182 (-0.024 to 0.388), 0.083 | | 77.85 | <.001 |  |
| **Femoral Neck** |  |  |  |  |  |  |
| Subtotal | 15 | 0.364 (0.134 to 0.595), 0.002 | | 71.46 | <.001 | 0.738 |
| Subtotal* | 13 | 0.249 (0.043 to 0.454), 0.018 | | 58.27 | 0.004 |  |
| **Total Hip** |  |  |  |  |  |  |
| Subtotal | 14 | 0.116 (-0.382 to 0.614), 0.648 | | 94.59 | <.001 | 0.747 |
| Subtotal* | 11 | -0.009 (-0.482 to 0.465), 0.971 | | 91.73 | <.001 |  |
| **Total Body** |  |  |  |  |  |  |
| Subtotal | 51 | 0.149 (0.006 to 0.291), 0.040 | | 80.84 | <.001 | 0.834 |
| Subtotal* | 29 | 0.199 (-0.022 to 0.420), 0.078 | | 85.58 | <.001 |  |

*The pooled results of sensitivity analyses excluding the studies of low or medium quality.
