## Supplementary file 5 for "The effect of calcium supplementation in people under 35 years old: A systematic review and meta-analysis of randomized controlled trials"

**Supplementary file 5A. Sensitivity analysis by comparisons of fixed and random effect models for bone mineral density (BMD)**

| **Subgroups** | **No. of studies** | **No. of participants** | | **Models** | **Effect estimates(95%CI)** | **P-Value** | **Heterogeneity between models** |
| --- | --- | --- | --- | --- | --- | --- | --- |
|  |  | **treatment group** | **control group** |  |  |  | **P-value** |
| **Total** | 54 | 3283 | 3459 | R | 0.413 (0.261 to 0.565) | <.001 | <.001 |
|  |  |  |  | F | 0.332 (0.277 to 0.386) | <.001 |  |
| **Lumbar Spine** | 35 | 1774 | 1824 | R | 0.090 (-0.044 to 0.224) | 0.190 | 0.806 |
|  |  |  |  | F | 0.059 (-0.010 to 0.129) | 0.094 |  |
| **Femoral Neck** | 24 | 1355 | 1054 | R | 0.627 (0.338 to 0.915) | <.001 | 0.007 |
|  |  |  |  | F | 0.391 (0.297 to 0.486) | <.001 |  |
| **Total Hip** | 18 | 866 | 910 | R | 0.257 (-0.053 to 0.566) | 0.104 | 0.641 |
|  |  |  |  | F | 0.152 (0.055 to 0.249) | 0.002 |  |
| **Total Body** | 38 | 1870 | 2013 | R | 0.330 (0.163 to 0.496) | <.001 | <.001 |
|  |  |  |  | F | 0.340 (0.278 to 0.403) | <.001 |  |

**Supplementary file 5B. Sensitivity analysis by comparisons of fixed and random effect models for bone mineral content (BMC)**

| **Subgroups** | **No. of studies** | **No. of participants** | | **Models** | **Effect estimates(95%CI)** | **P-Value** | **Heterogeneity between models** |
| --- | --- | --- | --- | --- | --- | --- | --- |
|  |  | **treatment group** | **control group** |  |  |  | **P-value** |
| **Total** | 55 | 2387 | 2522 | R | 0.285 (0.154 to 0.415) | <.001 | 0.002 |
|  |  |  |  | F | 0.254 (0.196 to 0.312) | <.001 |  |
|  |  |  |  | F | 0.224 (0.112 to 0.336) | <.001 |  |
| **Lumbar Spine** | 36 | 1331 | 1423 | R | 0.163 (0.008 to 0.317) | 0.039 | 0.324 |
|  |  |  |  | F | 0.136 (0.059 to 0.213) | 0.001 |  |
|  |  |  |  | F | 0.107 (-0.062 to 0.276) | 0.213 |  |
| **Femoral Neck** | 15 | 587 | 631 | R | 0.364 (0.134 to 0.595) | 0.002 | 0.135 |
|  |  |  |  | F | 0.318 (0.199 to 0.438) | 0.000 |  |
| **Total Hip** | 14 | 673 | 642 | R | 0.116 (-0.382 to 0.614) | 0.648 | 0.250 |
|  |  |  |  | F | 0.016 (-0.098 to 0.13) | 0.782 |  |
| **Total Body** | 51 | 2129 | 2265 | R | 0.149 (0.006 to 0.291) | 0.040 | 0.024 |
|  |  |  |  | F | 0.163 (0.102 to 0.224) | <.001 |  |
