## Supplementary file 6 for "The effect of calcium supplementation in people under 35 years old: A systematic review and meta-analysis of randomized controlled trials"

**Supplementary file 6A. Cumulative meta-analysis according to sample size in lumbar spine bone mineral density (LSBMD)**

**Supplementary file 6B. Cumulative meta-analysis according to sample size in femoral neck bone mineral density (FNBMD)**

**Supplementary file 6C. Cumulative meta-analysis according to sample size in total hip bone mineral density (THBMD)**

**Supplementary file 6D. Cumulative meta-analysis according to sample size in total body bone mineral density (TBBMD)**

**Supplementary file 6E. Cumulative meta-analysis according to sample size in lumbar spine bone mineral content (LSBMC)**

**Supplementary file 6F. Cumulative meta-analysis according to sample size in femoral neck bone mineral content (FNBMC)**

**Supplementary file 6G. Cumulative meta-analysis according to sample size in total hip bone mineral content (THBMC)**

**Supplementary file 6H. Cumulative meta-analysis according to sample size in total body bone mineral content (TBBMC)**
