## Supplementary file 7 for "The effect of calcium supplementation in people under 35 years old: A systematic review and meta-analysis of randomized controlled trials"

**Supplementary file 7A. Sensitivity analyses by comparisons of the pooled results of the trials included in previous study and trials newly-added in our current study of bone mineral density (BMD)***

| **Subgroups** | **No. of studies or subgroups** | **Effect estimate (95%CI)** | **P-value** | **Heterogeneity between studies** | | **P-value for heterogeneity between subgroups** |
| --- | --- | --- | --- | --- | --- | --- |
|  |  |  |  | **I² (%)** | **P-value** |  |
| **Lumbar Spine** |  | | | | |  |
| Trials included in previous study | 9 | 0.143 (-0.113 to 0.339) | 0.274 | 74.12 | <.001 | 0.673 |
| Trials newly included | 26 | 0.071 (-0.091 to 0.232) | 0.39 | 72.06 | <.001 |  |
| **Femoral Neck** |  | | | | |  |
| Trials included in previous study | 7 | 0.637 (0.003 to 1.271) | 0.049 | 93.86 | <.001 | 0.313 |
| Trials newly included | 17 | 0.603 (0.286 to 0.92) | <.001 | 83.49 | <.001 |  |
| **Total Hip** |  | | | | |  |
| Trials included in previous study | 1 | 0.683 (0.214 to 1.151) | 0.004 | 0.00 | 1.000 | 0.014 |
| Trials newly included | 17 | 0.232 (-0.089 to 0.553) | 0.157 | 89.98 | <.001 |  |
| **Total Body** |  | | | | | |
| Trials included in previous study | 7 | 0.171 (0.036 to 0.307) | 0.013 | 0.00 | 0.935 | 0.507 |
| Trials newly included | 31 | 0.365 (0.161 to 0.569) | <.001 | 87.49 | <.001 |  |

*The previous study are mentioned in the discussion part of our manuscript and are from Winzenberg, T et al.

**Supplementary file 7B. Sensitivity analyses by comparisons of the pooled results of the trials included in previous study and trials newly-added in our current study of bone mineral content (BMC)***

| **Subgroups** | **No. of studies or subgroups** | **Effect estimate (95%CI)** | **P-value** | | | **Heterogeneity between studies** | | **P-value for heterogeneity between subgroups** |
| --- | --- | --- | --- | --- | --- | --- | --- | --- |
|  |  |  |  |  |  | **I² (%)** | **P-value** |  |
| **Lumbar Spine** |  |  | |  |  |  |  |  |
| Trials included in previous study | 10 | 0.144 (-0.065 to 0.353) | 0.177 | | | 42.93 | 0.072 | 0.773 |
| Trials newly included | 26 | 0.174 (-0.024 to 0.373) | 0.086 | | | 78.69 | <.001 |  |
| **Femoral Neck** |  |  | |  |  |  |  |  |
| Trials included in previous study | 4 | 0.348 (-0.097 to 0.792) | 0.125 | | | 63.67 | 0.041 | 0.930 |
| Trials newly included | 11 | 0.369 (0.088 to 0.649) | 0.010 | | | 75.21 | <.001 |  |
| **Total Hip** |  |  | |  |  |  |  |  |
| Trials included in previous study | -- | -- | -- | | | -- | -- | -- |
| Trials newly included | 14 | 0.116 (-0.382 to 0.614) | 0.648 | | | 94.59 | <.001 |  |
| **Total Body** |  |  | |  |  |  |  |  |
| Trials included in previous study | 15 | 0.100 (-0.073 to 0.273) | 0.259 | | | 48.558 | 0.018 | 0.994 |
| Trials newly included | 36 | 0.177 (-0.009 to 0.363) | 0.063 | | | 84.986 | <.001 |  |

*The previous study are mentioned in the discussion part of our manuscript and are from Winzenberg, T et al.
