## Supplementary file 8 for "The effect of calcium supplementation in people under 35 years old: A systematic review and meta-analysis of randomized controlled trials"

**Supplementary file 8: GRADE assessment**

| **Certainty assessment** | | | | | | | **№ of patients** | | **Effect** | | **Certainty** | **Importance** |
| --- | --- | --- | --- | --- | --- | --- | --- | --- | --- | --- | --- | --- |
| **№ of studies** | **Study design** | **Risk of bias** | **Inconsistency** | **Indirectness** | **Imprecision** | **Other considerations** | **[干预]** | **[对照]** | **Relative (95% CI)** | **Absolute (95% CI)** |  |  |
| **LSBMD (follow-up: range 3 months to 2 years; assessed with: DXA)** | | | | | | | | | | | | |
| 35 | randomised trials | not serious | serious^a^ | not serious | not serious | none | 1774 | 1824 | - | SMD **0.09 SD higher** (-0.044 lower to 0.224 higher) | ⨁⨁⨁◯ Moderate | CRITICAL |
| **FNBMD (follow-up: range 9 months to 2 years; assessed with: DXA)** | | | | | | | | | | | | |
| 24 | randomised trials | not serious | serious^a^ | not serious | not serious | publication bias strongly suspected^b^ strong association^c^ | 1355 | 1054 | - | SMD **0.627 SD more** (0.338 more to 0.915 more) | ⨁⨁⨁◯ Moderate | CRITICAL |
| **THBMD (follow-up: range 8 months to 2 years; assessed with: DXA)** | | | | | | | | | | | | |
| 18 | randomised trials | not serious | serious^a^ | not serious | not serious^b^ | none | 866 | 910 | - | SMD **0.257 SD higher** (-0.053 lower to 0.566 higher) | ⨁⨁⨁◯ Moderate | CRITICAL |
| **TBBMD (follow-up: range 3 months to 2 years; assessed with: DXA)** | | | | | | | | | | | | |
| 38 | randomised trials | not serious | serious^a^ | not serious | not serious | none | 1870 | 2013 | - | SMD **0.33 SD more** (0.163 more to 0.496 more) | ⨁⨁⨁◯ Moderate | CRITICAL |
| **LSBMC (follow-up: range 3 months to 2 years; assessed with: DXA)** | | | | | | | | | | | | |
| 36 | randomised trials | not serious | serious^a^ | not serious | not serious | none | 1331 | 1423 | - | SMD **0.163 SD more** (0.008 more to 0.317 more) | ⨁⨁⨁◯ Moderate | CRITICAL |
| **FNBMC (follow-up: range 1 years to 2 years; assessed with: DXA)** | | | | | | | | | | | | |
| 15 | randomised trials | not serious | serious^a^ | not serious | not serious^b^ | none | 587 | 631 | - | SMD **0.364 SD more** (0.134 more to 0.595 more) | ⨁⨁⨁◯ Moderate | CRITICAL |
| **THBMC (follow-up: range 1 years to 2 years; assessed with: DXA)** | | | | | | | | | | | | |
| 14 | randomised trials | not serious | serious^a^ | not serious | not serious^b^ | none | 673 | 642 | - | SMD **0.116 SD higher** (-0.382 lower to 0.614 higher) | ⨁⨁⨁◯ Moderate | CRITICAL |

**eTable 8: GRADE assessment (continued)**

| **TBBMC (follow-up: range 3 months to 2 years; assessed with: DXA)** | | | | | | | | | | | | |
| --- | --- | --- | --- | --- | --- | --- | --- | --- | --- | --- | --- | --- |
| 51 | randomised trials | not serious | serious^a^ | not serious | not serious | none | 2129 | 2265 | - | SMD **0.149 SD more** (0.006 more to 0.291 more) | ⨁⨁⨁◯ Moderate | CRITICAL |

Abbreciations: CI: confidence interval; SMD: standardised mean difference

a. We conducted I2 test for heterogeneity, all studies included were over 60% and had some heterogeneity

b. We assessed publication bias by examining funnel plots and using Begg’s rank correlation and Egger’s linear regression tests, and this outcome was strongly suspected.

c. When effect size was over 0.5, it was determined as upgrade +1.
