## Supplementary file 9 for "The effect of calcium supplementation in people under 35 years old: A systematic review and meta-analysis of randomized controlled trials"

**Supplementary file 9: Meta-regression for age, region, Ca dosage, baseline intake and sample size on bone mineral density (BMD) and bone mineral content (BMC)**

| **Covariate** | | **Coefficient** | **Standard Error** | **95% Lower** | **95% Upper** | **Z-Value** | **2-sided P-value** |
| --- | --- | --- | --- | --- | --- | --- | --- |
| **Intercept** | BMD | 0.3116 | 0.5339 | -0.7347 | 1.358 | 0.58 | 0.5594 |
|  | BMC | 0.3078 | 0.3402 | -0.359 | 0.9745 | 0.9 | 0.3656 |
| **Age** | BMD | -0.0528 | 0.0288 | -0.1093 | 0.0037 | -1.83 | 0.0668 |
|  | BMC | -0.0142 | 0.0178 | -0.0491 | 0.0207 | -0.8 | 0.4247 |
| **Region** | BMD | 1.1614 | 0.2922 | 0.5888 | 1.7341 | 3.98 | 0.0001 |
|  | BMC | 0.3098 | 0.1946 | -0.0715 | 0.6911 | 1.59 | 0.1113 |
| **Ca dosage** | BMD | -0.0006 | 0.0004 | -0.0014 | 0.0002 | -1.39 | 0.1639 |
|  | BMC | 0.0001 | 0.0002 | -0.0003 | 0.0006 | 0.6 | 0.5516 |
| **Baseline intake** | BMD | -0.0009 | 0.0007 | -0.0023 | 0.0004 | -1.39 | 0.1636 |
|  | BMC | -0.0007 | 0.0004 | -0.0014 | 0.0001 | -1.68 | 0.0922 |
| **Sample size** | BMD | 0.0009 | 0.0008 | -0.0007 | 0.0025 | 1.06 | 0.2905 |
|  | BMC | 0.0000 | 0.0007 | -0.0013 | 0.0013 | 0.02 | 0.9845 |
