## Supplementary file 10 for "The effect of calcium supplementation in people under 35 years old: A systematic review and meta-analysis of randomized controlled trials"

**Supplementary file 10. Publication bias**

| **Subgroups( by sites)** | **No. of datasets** | **P-Value for** | | **Effect estimates (95%CI)*** | |
| --- | --- | --- | --- | --- | --- |
|  |  | **Begg's rank correlation test** | **Egger's linear regression test** | **Adjusted with trim and fill** | **Unajusted** |
| **BMD** | | | | | |
| Lumbar spine | 35 | 0.514 | 0.197 | 0.090 (-0.044, 0.224) | 0.090 (-0.044, 0.224) |
| Femoral neck | 24 | 0.007 | 0.006 | 0.331 (-0.0005, 0.663) | 0.627 (0.338, 0.915) |
| Total hip | 18 | 0.080 | 0.188 | -0.118 (-0.461, 0.226) | 0.257(-0.053, 0.566) |
| Total body | 38 | 0.497 | 0.760 | 0.330 (0.163, 0.496) | 0.330 (0.163, 0.496) |
| **BMC** | | | | | |
| Lumbar spine | 36 | 0.859 | 0.369 | 0.163 (0.008, 0.317) | 0.163 (0.008, 0.317) |
| Femoral neck | 15 | 0.553 | 0.271 | 0.253 (0.003, 0.502) | 0.364 (0.134, 0.595) |
| Total hip | 14 | 0.004 | 0.489 | 0.116 (-0.382, 0.614) | 0.116 (-0.382, 0.614) |
| Total body | 51 | 0.16 | 0.65 | 0.149 (0.006, 0.291) | 0.149 (0.006, 0.291) |

Abbreviations: BMD, bone mineral density; BMC, bone mineral content; CI, confidence interval.

*The summarized effect estimate was calculated with a random-effects model.
